# Systematic evaluation of plasma and urine metabolites to predict adverse kidney-related outcomes in chronic kidney disease: The GCKD study

**DOI:** 10.1101/2025.05.09.25326983

**Authors:** Elena Butz, Inga Steinbrenner, Ulla T. Schultheiss, Charlotte Behning, Harald Binder, Helena Hansmann, Wolfram Gronwald, Peter J. Oefner, Elke Schaeffner, Kai-Uwe Eckardt, Anna Köttgen, Peggy Sekula, GCKD Investigators

## Abstract

**Rationale & Objective:** Accurate risk prediction of adverse kidney-related outcomes in individuals with chronic kidney disease (CKD) is essential to guide personalized clinical interventions. Metabolites measured in plasma and urine may offer additional prognostic information beyond established clinical predictors.

**Study Design:** Prospective German CKD cohort study.

**Setting & Participants:** 5,217 individuals with predominantly CKD stage G3 at baseline, followed for a median of 6.5 years (IQR 6.5-6.5).

**Exposure(s) *or* • Predictor(s):** Baseline metabolite levels measured via untargeted mass spectrometry (Metabolon Inc.) in plasma (N=5,144; 1,096 metabolites) and urine (N=5,088; 1,129 metabolites).

**Outcome(s):** (i) Kidney failure (KF), defined as initiation of maintenance dialysis, kidney transplantation, or death due to untreated KF; (ii) composite kidney endpoint (CKE), defined as KF, a ≥40% decline in estimated glomerular filtration rate (eGFR), or an eGFR <15 mL/min/1.73 m².

**Analytical Approach:** Time-to-event data were analyzed using subdistribution hazard models to predict outcomes. A component-wise boosting algorithm was employed to select metabolites in various scenarios, including models based on plasma, urine, or combined data. Predictive performance was assessed at multiple time points using time-dependent area under the curve (AUC) and Brier scores, and compared to benchmark models.

**Results:** Several individual metabolites were predictive of KF or CKE and provided marginal improvements beyond established prognostic factors (age, sex, eGFR, and urinary albumin-to-creatinine ratio [UACR]). For example, plasma pseudouridine increased the AUC at year 6 by 0.012 (95% CI: 0.005–0.018) when added to the clinical model.

Multi-metabolite models developed under different scenarios included a median of 53 metabolites (range: 14-86). Some metabolites, such as plasma N2,N5-diacetylornithine and urine 1-palmitoyl-2-oleoyl-GPC (16:0/18:1), were selected more often than others. Predictive performance of all models declined over time. For KF, models achieved AUCs ≥0.89 at year 2 and ≥0.85 at year 6; for CKE, AUCs were ≥0.85 and ≥0.77, respectively. Compared to benchmark models with only clinical variables, metabolite-based models performed better but not to a clinically meaningful extent. AUC values were comparable to those reported for other published KF prediction models.

**Limitations:** Use of single baseline measurements and semi-quantitative nature of the metabolite measurements.

**Conclusions:** While certain metabolites improved prediction of adverse kidney-related outcomes, the observed gains were not clinically meaningful. Nonetheless, these findings may provide insights into metabolic pathways and processes related to the progression of CKD, thereby complementing current knowledge. Further research is warranted to refine predictive models and explore the biological relevance of identified metabolites.

## Introduction

Disease courses of individuals with chronic kidney disease (**CKD**) are highly variable^1,2^. It is thus clinically important to identify individuals at higher risk for adverse outcomes such as kidney failure (**KF**), to enable earlier and more personalized interventions^3–5^.

Several prognostic models for KF have been developed, with the most widely used being the kidney failure risk equation (**KFRE**)^6^ containing the four variables age, sex, When assessing its performance across multiple studies, the authors found that the model distinguished well between individuals experiencing KF and those who did not (C-index at 5 years: 0.88, 95% confidence interval [CI]: 0.86–0.90)^8^. However, they also noted some variability in performance across studies, with C-index values between 0.77 and 0.96.

Attempts to improve KF risk prediction with additional clinical parameters have shown limited success^6,9–12^.

Metabolites are promising prognostic biomarkers in nephrology because they reflect (patho-)physiological processes and are often cleared by the kidneys^13,14^. Several studies, including our own work within the German Chronic Kidney Disease (**GCKD**) study, have examined associations between metabolite levels and adverse CKD outcomes^14–17^. Recently, we showed that certain individual metabolites can improve the prediction of adverse kidney outcomes beyond the four KFRE variables^17^.

These findings prompted us to extend the previous assessment to a systematic evaluation of the prognostic value of all metabolites measured in baseline plasma and urine samples from participants in the GCKD study^18^. In addition to evaluating individual metabolites, we used a feature selection algorithm to develop multi-metabolite models, combining prognostic values of single metabolites. We then compared the predictive performance of these models to benchmark models, including the KFRE, to determine their added value in predicting adverse kidney-related outcomes^7,19^.

## Methods

### Study population

The GCKD study is an ongoing multicenter prospective cohort study (DRKS 00003971) that enrolled 5,217 participants with CKD between 2010 and 2012^18^. Major inclusion criteria were (a) an eGFR between 30 and 60 mL/min/1.73m², or (b) overt albuminuria or proteinuria with an eGFR >60 mL/min/1.73m². All participants had been referred to nephrologists and provided informed consent. The local ethics committees of the participating centers approved the study (**Item S1**).

### Baseline data and measurements

At baseline, participants were systematically assessed by trained study personnel to collect demographic and clinical data. Biospecimens, including plasma and spot urine, were obtained and transported frozen to a central biobank, following standardized protocols^20^. Some laboratory parameters were measured immediately, while other samples were stored for future analysis. Details of the study cohort have been published previously^21^. Variables used in this analysis are listed in **Item S2**. eGFR was calculated using the CKD-EPI creatinine-based equation^22^, consistent with our prior work^17^.

Baseline urine and plasma samples were analyzed by Metabolon Inc. (Durham, NC, USA) using untargeted liquid chromatography-mass spectrometry. Urine samples were processed in three batches between 2016 and 2017, and plasma samples in a single batch in 2020. Details on sample preparation, MS procedures, and metabolite identification have been described previously^17,23^, with a summary provided in **Item S3**. Named metabolites were assigned to super-pathways (e.g., amino acids, xenobiotics). Unless marked with an asterisk, named metabolites met the highest confidence level (Level 1) per Metabolomics Standards Initiative criteria^24,25^. Some metabolites were partially characterized or unnamed, lacking a defined structural identity.

Following release of the semi-quantitative data, measurements underwent quality control and preprocessing (**Item S4**). Missing values were imputed using the k-nearest neighbor (*knn*) algorithm^26^, enabling the application of feature selection methods for developing multi-metabolite models. Imputation was restricted to metabolites with sufficient metabolites (609 named) were excluded due to high proportions of missing data. Quality assessment of the imputed data did not indicate any issues of concern (see **Item S5** for details).

The final dataset included measurements of 1,096 plasma metabolites from 5,144 individuals and 1,129 urine metabolites from 5,088 individuals (see **Figure S1** and **Tables S1–S2**). Of the included metabolites, 25% in plasma and 38% in urine were unnamed. A total of 583 metabolites (417 named) were detected in both plasma and urine. Paired plasma and urine measurements were available for 5,023 participants.

### Prospective Data and Definition of Outcomes

Events were identified using hospital discharge summaries, nephrologist outpatient reports and death certificates, and were adjudicated by a medical expert team using a standardized event catalog.^27^ In line with our previous work^17^, follow-up data up to 6.5 years after study enrollment were included in the analysis (data freeze: March 31, 2021).

The primary outcomes were KF, defined as the initiation of kidney replacement therapy (maintenance dialysis or kidney transplantation) or death due to untreated KF, and a composite kidney endpoint (**CKE**), which included KF, a ≥40% decline in eGFR, or an eGFR <15 mL/min/1.73m², both based on estimated eGFR slopes.

### Statistical Analysis

The main workflow is displayed in **Figure S1**. All analyses were performed using R (version 4.0.5 or 4.3.3, R Core Team (2020) – R: A language and environment for statistical computing. R Foundation for Statistical Computing, Vienna, Austria, http://www.R-project.org/). This project involved two primary steps: First, evaluating the ability of individual metabolites to predict KF and CKE, and second, developing multi-metabolite models for various settings and evaluating their predictive performance (**Item S6**).

*(i) Regression models:* Subdistribution hazard models were chosen to predict the risk of outcomes, since they account for competing risks such as death from causes other than untreated KF^28,29^. Outcomes were defined as the time from study entry to either the event of interest or a competing event, whichever occurred first. Individuals who experienced neither were censored at their last known time alive within the period of maximum 6.5 years.

To assess the prognostic value of metabolites alone and in addition to known prognostic factors, we evaluated three model configurations:

- MET: metabolites only,
- MET+: metabolites plus KFRE variables (age, sex, eGFR, and ln-transformed UACR)^7^, and
- MET++: Metabolites plus an extended set of 14 clinical prognostic factors (see **Item S2**). The extended set corresponds to the comprehensive model used in our previous project^17^ and includes variables selected based on a thorough literature review and biological rationale to encompass additional risk and prognostic factors (Status: August 2021).
*(ii) Handling of missing observations:* Due to the imputation of missing measurements, metabolite data were complete for individuals with any metabolites available in the respective matrix (plasma or urine, **Item S4**). The proportion of missing data for clinical prognostic factors ranged from 0% to 2% (**Item S2**).

For models involving one single metabolite (Step 1), we used two subsets of the GCKD study population that were restricted to individuals with complete data on the four KFRE variables and respective metabolite measurements in plasma (N=5,047) or urine (N=5,025; **Figure S1**). For Step 2 (multi-metabolite models), we included only individuals with complete data for all 14 prognostic factors and the relevant metabolite measurements (**Item S2**). These subsets included 4,947 individuals with plasma measurements, 4,925 with restriction ensured comparability of the predictive performances across different model setups for a specific matrix.

*(iii) Multi-metabolite model construction:* To select metabolites and to fit multi-metabolite models, the CoxBoost (v1.5; https://github.com/binderh/CoxBoost) algorithm, a component-wise boosting algorithm for subdistribution hazard models, was employed^30^. The algorithm performs a stepwise update of the coefficient of one variable at a time using penalized partial likelihood estimation. Prior to metabolite selection, the number of pre-specified boosting steps for each setting was determined using 10-fold cross-validation. Only metabolites were subjected to feature selection, while clinical predictors were kept mandatory, if present in the model. Detailed descriptions of the selected parameters are provided in **Item S7**.

The selection process was repeated for metabolites from each matrix separately (plasma or urine) as well as for the combined matrix set (see **Item S6**). In total, 18 model settings were evaluated (2 outcomes × 3 matrix sets × 3 model configurations). Initially, only metabolites named by Metabolon were included in the feature selection. Subsequently, all 18 settings were re-run including unnamed metabolites to assess whether they provided additional prognostic information beyond the named ones.

To study the stability of the selection of metabolites by CoxBoost, we generated 100 random subsamples, each comprising 63.2% of the analysis set without replacement, and observed how frequently metabolites were selected across the subsamples^31^. The entire procedure, including determination of the required boosting steps, was repeated for each subsample.

*(iv) Assessment of predictive performance:* To comprehensively assess the predictive performance of each model, we used the R package *riskRegression* (version 2023.01.19; https://github.com/tagteam/riskRegression) developed for regression models with survival outcomes in the presence of competing risks^32,33^. To prepare *CoxBoost* objects to be assessed with *riskRegression*, we used a wrapper function to uniformly revise these objects.

Discrimination between individuals with and without the event of interest was assessed using time-dependent area under the receiver operating characteristic curve (**AUC**) and evaluated at 2, 4, 5, and 6 years after study entry^33,34^. The prediction error (Brier score) was assessed at the same time points^35^. For both measures, estimates were also obtained using bootstrap cross-validation^19^. When not indicated otherwise, reported values correspond to bootstrap cross-validated results.

The predictive performance of each multi-metabolite model was compared to its respective benchmark model, which corresponded to the same model without metabolites: MET+ vs **Bench+** (comprising the four KFRE variables), MET++ vs **Bench++** (comprising the extended set of 14 prognostic factors; **Item S2)**. For further comparison, we applied other published proposals of models predicting KF to our analysis data^9,10,36^. Notably, the model proposed by Zacharias *et al*.^36^ was also developed using GCKD study data, albeit with a shorter follow-up period of 4 years.

The reporting guideline REMARK for prognostic factor studies was considered when drafting the manuscript^37^.

## Results

### Study population and metabolite data

Depending on the availability of plasma and/or urine metabolite measurements and completeness of data on prognostic variables, the analysis cohorts varied slightly in size (**Figure S1**).

Baseline characteristics of individuals across all analysis cohorts (N = 4,886 – 5,047) were comparable to those of the full GCKD study population (N=5,217; **Table S3**). The proportion of males was 60%, the mean age was 60 years, mean eGFR was 49.5 mL/min/1.73m², and the median UACR ranged between 50.4 and 50.7 mg/g (**Table 1**). Prevalent morbidities at baseline included hypertension (96%), diabetes (35%), coronary heart disease (20%), and stroke (10%) (**Table S3**).

**Table 1:** Overview of sample characteristics for different analysis sets.

| Variable | GCKD cohort | Step 1: evaluation of individual metabolites<sup>1</sup> |  | Step 2: evaluation of multi-metabolite models<sup>2</sup> |  |  |
| --- | --- | --- | --- | --- | --- | --- |
|  |  | cohort with plasma metabolites | cohort with urine metabolites | cohort with plasma metabolites | cohort with urine metabolites | cohort with plasma and urine metabolites |
| Sample size | 5,217 | 5,047 | 5,025 | 4,947 | 4,925 | 4,886 |
| Sex, male | 60 (3132) | 60 (3035) | 60 (3025) | 60 (2980) | 60 (2971) | 60 (2947) |
| Age (years) | 60.1 (12.0) | 60.0 (12.0) | 60.0 (12.0) | 60.0 (12.0) | 60.0 (12.0) | 60.0 (12.0) |
| eGFR (mL/min/1.73m<sup>2</sup>) | 49.4 (18.3) | 49.5 (18.3) | 49.5 (18.3) | 49.5 (18.3) | 49.5 (18.3) | 49.5 (18.3) |
| UACR (mg/g) | 50.9<br>(9.7,392) | 50.7<br>(9.6,391) | 50.7<br>(9.6,387) | 50.7<br>(9.5,388) | 50.7<br>(9.6,383) | 50.4<br>(9.5,385) |

1: restriction to cases with measurements of respective matrix and complete information on age, sex, eGFR and UACR; 2: restriction to cases with measurements of respective matrix and complete information on extended list of prognostic factors; see **Item S2** for details on prognostic factors included.
For age and eGFR, mean (SD) is provided and median (IQR) for UACR. For 55 and 90 individuals, values of eGFR or UACR, respectively, are missing.
Abbreviations: eGFR, estimated glomerular filtration rate; UACR, urinary albumin-to-creatinine ratio.

Over a median follow-up of 6.5 years (IQR 6.5-6.5), the proportions of incident KF events (Nevents=500, 10%) and CKE (Nevents=1,083, 21%) observed in the full GCKD study cohort (N = 5,217) was mirrored in all analysis cohorts. Among the 1,083 CKE cases, 832 were attributable to a ≥40% eGFR decline, 226 to an eGFR <15 mL/min/1.73m², and 142 to KF. All KF cases are also CKE cases; however, in 358 of 500 individuals with KF, a CKE event occurred a median 510 days earlier than KF due to early occurrence of ≥40% eGFR decline or reaching the <15 mL/min/1.73m² threshold.

### Prognostic value of individual metabolites

In the first step, we evaluated the ability of each metabolite to predict KF and CKE individually (MET model), as well as its added prognostic value beyond the four KFRE variables (age, sex, eGFR, and ln-transformed UACR) by comparing MET+ to Bench+ models.

Several metabolites showed strong discriminatory power when used alone (MET model). Among plasma metabolites, pseudouridine achieved the highest observed AUC for predicting KF at year 6 (AUC: 0.80, 95% CI: 0.77–0.83; **Table S4**). For comparison, plasma creatinine (measured by Metabolon) had an AUC of 0.78 (95% CI: 0.74-0.81). For CKE, the top-performing plasma metabolite was the unnamed metabolite X-25422 with AUC of 0.72 (95% CI 0.70-0.74), followed by pseudouridine (AUC: 0.71, 95% CI 0.68-0.73; **Table S5**). In urine, choline phosphate was the most predictive metabolite for both KF (AUC: 0.73, 95% CI: 0.70–0.76) and CKE (AUC: 0.68, 95% CI 0.66-0.71; **Tables S6-S7**).

The predictive performance of individual metabolites, as measured by AUC, generally declined over time. For instance, the AUC of plasma pseudouridine for KF prediction decreased from 0.88 at year 2 (95% CI: 0.84–0.91) to 0.81 at year 5 (95% CI: 0.78–0.84).

Adding individual metabolites to the Bench+ model rarely resulted in substantial improvements in AUC, suggesting limited added prognostic value in most cases (**Figures S2- S3**). Nonetheless, depending on the outcome and matrix, between 13 and 70 metabolites showed a statistically significant improvement in AUC at year 6 (**Tables S4–S7**), visible as upward outliers in AUC difference plots and downward outliers in Brier score difference plots (**Figures S2–S3**). For example, inclusion of plasma pseudouridine in the KF model improved the AUC at year 6 to 0.86 (95% CI: 0.84–0.88), representing a gain of 0.012 (95% CI: 0.005– 0.018) over the Bench+ model.

### Composition of multi-metabolite models

Multi-metabolite models were derived for a total of 36 distinct settings: 18 using only named metabolites, and 18 including both named and unnamed metabolites. In each case, the optimal number of boosting steps was determined before the selection started (**Item S7, Table S8**).

For instance, to develop a MET model predicting KF using only named plasma metabolites, 322 boosting steps were determined and yielded 50 selected metabolites. The coefficient trajectories of selected metabolites across boosting steps are illustrated in **Figure S4**, where the coefficient of a single metabolite is updated at each step.

Overall, the CoxBoost algorithm performed a median of 233.5 boosting steps (range: 34–499), resulting in a median of 53 metabolites selected per model (range: 14–86; **Table S8**). The number of selected metabolites increased with the number of boosting steps (Pearson r = 0.86). Models predicting CKE generally required more boosting steps and selected more metabolites than those predicting KF.

Across model configurations, the number of boosting steps and selected metabolites was highest in MET models (mean: 316.3 steps; 68.4 metabolites) and lowest in MET++ models (mean: 156.8 steps; 39.6 metabolites). This pattern suggests that some metabolites may be considered as potential proxies for clinical variables, while others provide additional prognostic value even in the presence of clinical information.

In the 18 settings limited to named metabolites, 271 distinct metabolites were selected at least once (mean: 45.4 per model; **Tables S8–S9**). Each metabolite could be selected in up to 12 settings.Among the selected metabolites, two stood out for their consistent inclusion: plasma N2,N5-diacetylornithine and urine 1-palmitoyl-2-oleoyl-GPC (16:0/18:1), each selected in all 12 relevant models (**Figure 1**). In total, seven plasma and eleven urine metabolites were selected in at least 8 out of 12 possible settings. As expected, the endogenous filtration marker creatinine (plasma, measured by Metabolon) was only selected in MET models predicting KF - i.e., models that, unlike MET+ and MET++ configurations, did not include eGFR.

**Figure 1:**
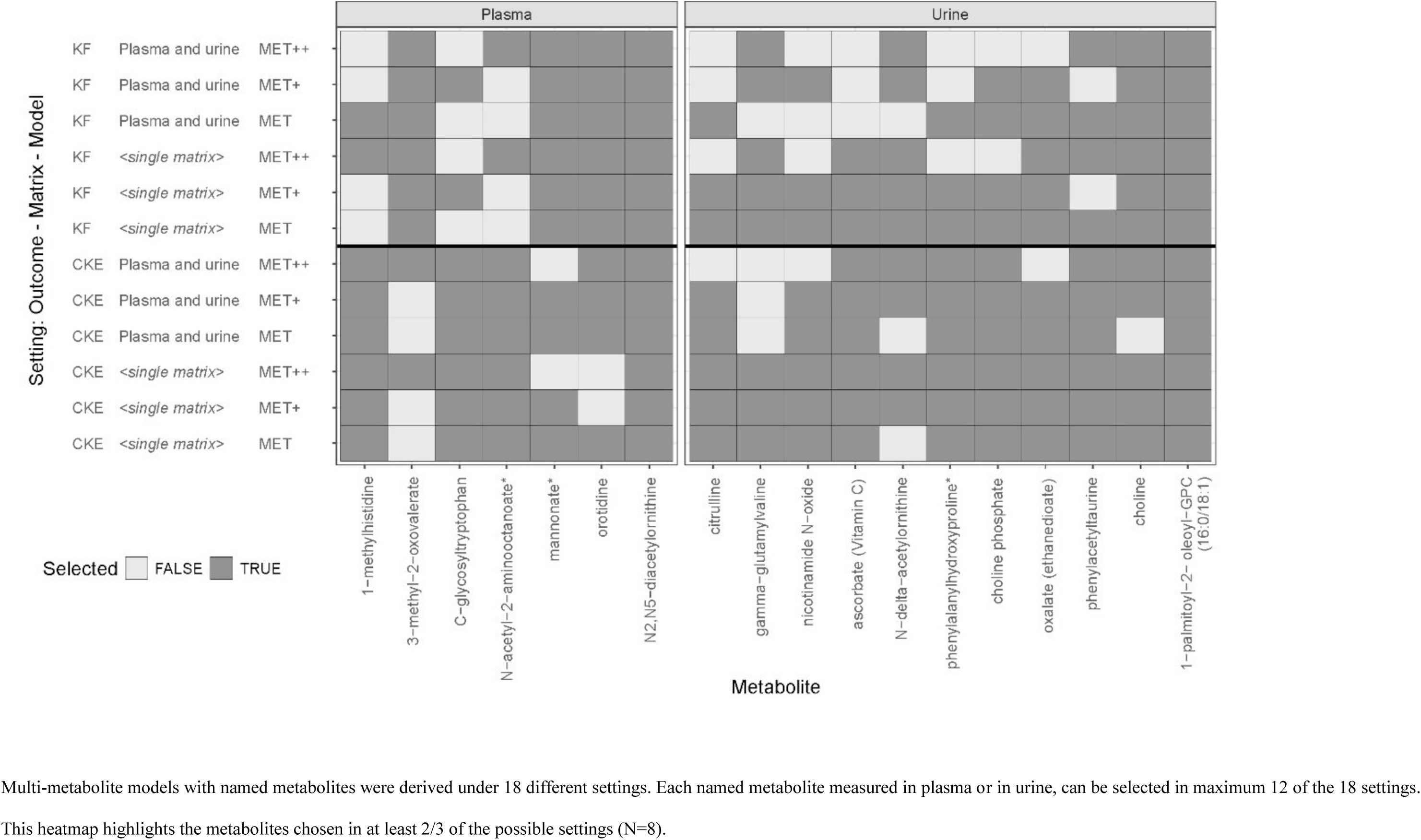
Overview of metabolites selected for at least 8 of 12 multi-metabolite models with named metabolites. Multi-metabolite models with named metabolites were derived under 18 different settings. Each named metabolite measured in plasma or in urine, can be selected in maximum 12 of the 18 settings. This heatmap highlights the metabolites chosen in at least 2/3 of the possible settings (N=8).

In the 18 settings where unnamed metabolites were also included in the selection process, the number of metabolites selected at least once increased to 304 (mean per model: 56.3). However, not all of the 271 named metabolites previously selected in the named-only settings were selected again (**Tables S8–S9**). Still, the two named metabolites that had been selected in all 12 applicable named-only models were again selected in 11 out of 12 models (**Figure S5**). Additionally, two unnamed plasma metabolites, X-24307 and X-25422, were selected in all 12 possible settings, highlighting the potential relevance of unnamed metabolites.

To evaluate the stability of metabolite selection by CoxBoost, we generated resampled datasets without replacement^31^. As these resampled datasets were smaller and included fewer events (**Table S10**), the median number of boosting steps and selected metabolites was frequently in the resampling procedure than those not originally selected (**Figure S6**). Fewer than half of the metabolites not selected in the main model appeared in any bootstrap sample, with the maximum frequency being 56 out of 100 subsamples (**Table S10**). In contrast, metabolites that had been selected in the original model were often reselected, with individual inclusion frequencies reaching up to 100% in MET models and up to 99% in the other models. The proportion of metabolites from the main analysis that were selected in at least 40 of the 100 subsamples ranged from 14.5% to 89.5% (median 46.5%).

### Prognostic value of multi-metabolite models

i. *Outcome KF:* All multi-metabolite models discriminated well between individuals who developed KF within 6 years and those who did not, with AUC values ≥0.85 (**Table 2**, **Figures S7–S8**). However, as seen for the benchmark models as well, performance declined over time in terms of both AUC and Brier score (**Tables S11 and S13, Figure 2**). For example, the MET++ model using named plasma and urine metabolites reached an AUC of 0.89 (95% CI 0.84–0.94) at year 2, which declined to 0.87 (95% CI 0.84–0.89) at year 5, with no further change at year 6.

Adding selected metabolites to the Bench+ model significantly increased AUCs. The greatest improvement was obtained by adding plasma und urine metabolites (including unnamed) with ΔAUC = 0.046 (95% CI 0.017-0.076) observed at year 2, 0.026 (95% CI 0.012-0.037) at year 5 and 0.022 (95% CI 0.011-0.033) at year 6 (**Table S14**). In contrast, when metabolites were added to the Bench++ model, improvements were smaller and no longer statistically significant at years 5 and 6. Reductions in prediction error (Brier score) were generally not statistically significant.

**Figure 2:**
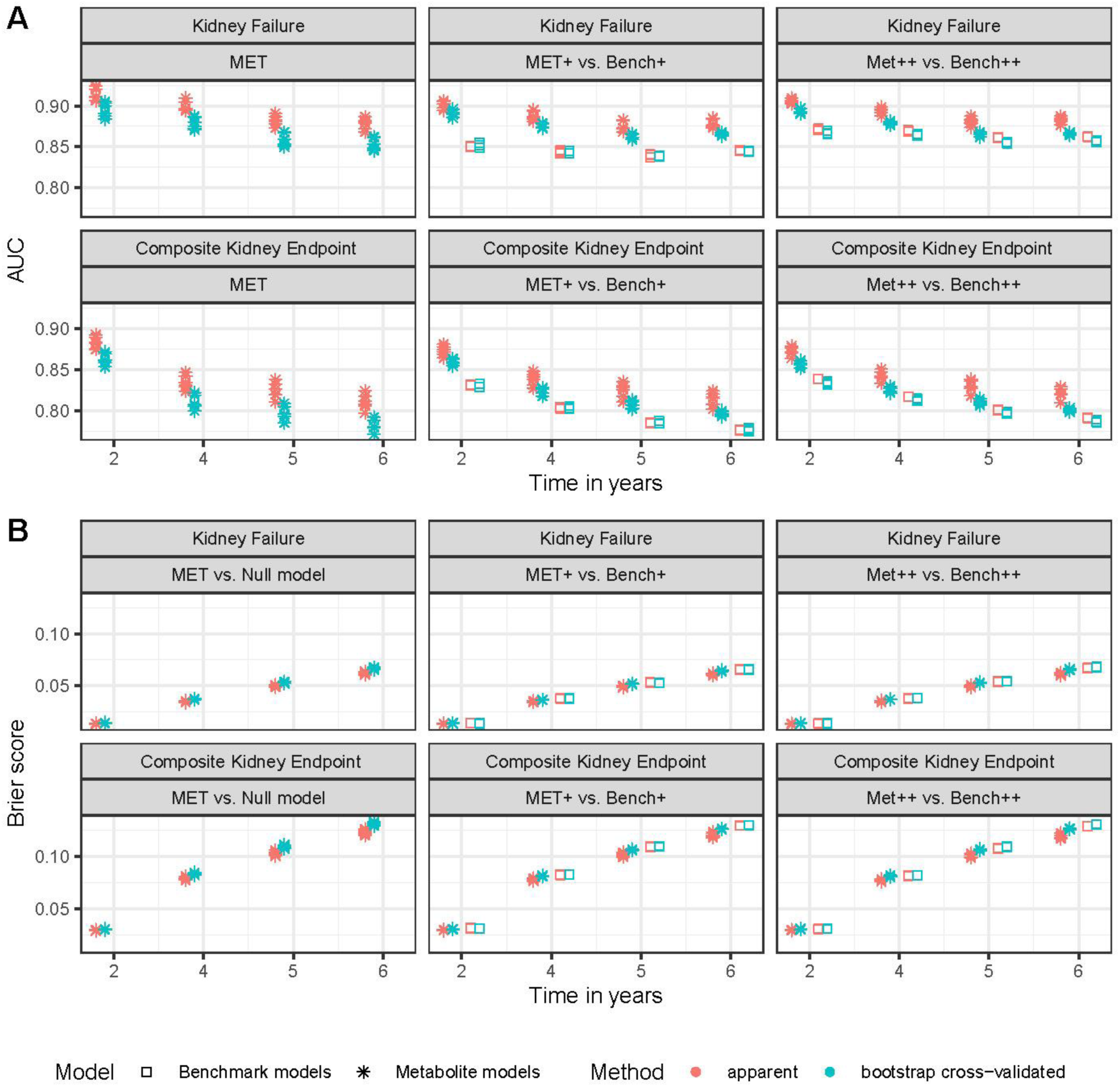
Performance of multi-metabolite models and benchmark models predicting kidney failure and composite kidney endpoint Displayed metrics illustrate the prognostic ability of models in terms of (A) AUC and (B) Brier score at years 2, 4, 5 and 6 for predicting kidney failure (upper row) and composite kidney endpoint (lower row). For AUC, higher values represent better discrimination of individuals with and without respective outcome. In contrast, lower Bier score (prediction error) reflects better prediction. Since apparent values of the predictive performance (in red) might be over-optimistic, bootstrap cross-validated values are in blue. For each time point and color, six stars are shown representing six different multi- metabolite models; models with all or only named metabolites (2) per matrix (3). For benchmark models, three squares representing each matrix are shown for each time point. In general, confidence intervals for AUC and Brier scores of multi- metabolite models do overlap with benchmark models, which is why they are not included here but can be looked up in **Tables S11-S13**. Confidence limits of calculated differences between multi-metabolite model and respective benchmark Abbreviations: AUC, area under the receiver operating characteristic curve; MET, multi-metabolite model w/o clinical information; MET+, multi-metabolite model including KFRE variables; MET++, multi-metabolite model including extended set of prognostic factors (see **Item S2** for details); Bench+/Bench++: model containing respective set of clinical information without metabolites; KFRE, kidney failure risk equation which contains age, sex, estimated glomerular filtration rate and urinary albumin-to-creatinine ratio

**Table 2:** Bootstrap cross-validated values of the area under ROC curve at <u>year 6</u> for prediction of kidney failure and composite kidney endpoint.

| Selection | Matrix | AUC (95% CI), bootstrap cross-validated |  |  |
| --- | --- | --- | --- | --- |
|  |  | Model: without clinical information (MET) | Model: including KFRE variables (MET+, Bench+) | Model: including extended set of prognostic factors (MET++, Bench++) |
| Outcome: kidney failure (KF) |  |  |  |  |
| Benchmark (w/o metabolites) | Plasma | NA | 0.84 (0.82-0.87) | 0.86 (0.83-0.88) |
| Based on named metabolites | Plasma | 0.85 (0.82-0.87) | 0.86 (0.84-0.88) | 0.87 (0.85-0.89) |
| Based on all metabolites | Plasma | 0.85 (0.83-0.87) | 0.87 (0.84-0.89) | 0.87 (0.85-0.89) |
| Benchmark (w/o metabolites) | Urine | NA | 0.85 (0.82-0.87) | 0.86 (0.83-0.88) |
| Based on named metabolites | Urine | 0.85 (0.82-0.87) | 0.86 (0.84-0.89) | 0.86 (0.84-0.89) |
| Based on all metabolites | Urine | 0.85 (0.82-0.87) | 0.86 (0.84-0.89) | 0.86 (0.84-0.89) |
| Benchmark (w/o metabolites) | Plasma, urine | NA | 0.84 (0.82-0.87) | 0.86 (0.83-0.88) |
| Based on named metabolites | Plasma, urine | 0.86 (0.84-0.88) | 0.86 (0.83-0.89) | 0.87 (0.84-0.89) |
| Based on all metabolites | Plasma, urine | 0.86 (0.84-0.88) | 0.87 (0.84-0.89) | 0.87 (0.84-0.89) |
| Outcome: composite kidney endpoint (CKE) |  |  |  |  |
| Benchmark (w/o metabolites) | Plasma | NA | 0.78 (0.76-0.80) | 0.79 (0.77-0.81) |
| based on named metabolites | Plasma | 0.77 (0.75-0.79) | 0.79 (0.78-0.81) | 0.80 (0.78-0.82) |
| based on all metabolites | Plasma | 0.78 (0.76-0.80) | 0.79 (0.77-0.81) | 0.80 (0.78-0.82) |
| Benchmark (w/o metabolites) | Urine | NA | 0.78 (0.76-0.80) | 0.79 (0.77-0.81) |
| based on named metabolites | Urine | 0.78 (0.76-0.80) | 0.80 (0.77-0.82) | 0.80 (0.78-0.82) |
| based on all metabolites | Urine | 0.78 (0.76-0.80) | 0.79 (0.77-0.82) | 0.80 (0.78-0.82) |
| Benchmark (w/o metabolites) | Plasma, urine | NA | 0.77 (0.75-0.80) | 0.79 (0.76-0.80) |
| based on named metabolites | Plasma, urine | 0.79 (0.77-0.81) | 0.80 (0.78-0.82) | 0.80 (0.78-0.82) |
| based on all metabolites | Plasma, urine | 0.79 (0.77-0.82) | 0.80 (0.78-0.82) | 0.80 (0.78-0.83) |

Abbreviations: AUC, area under the receiver operating characteristic curve; 95% CI, 95% confidence interval; KFRE, kidney failure risk equation which contains age, sex, estimated glomerular filtration rate and urinary albumin-to-creatinine ratio; MET, multi-metabolite model without clinical information; MET+, multi-metabolite model including KFRE variables; MET++, multi-metabolite model including extended set of prognostic factors (see **Item S2** for details); Bench+/Bench++: model containing respective set of clinical information without metabolites; NA, not applicable.

When we applied previously published models^9,10,36^ to our data, their performance in predicting KF (AUC range: 0.84–0.88) was comparable to our multi-metabolite models,

(ii) *Outcome CKE:* Compared to KF, multi-metabolite models for CKE showed acceptable but lower predictive performance, with AUC values declining over time (**Tables S12 and S13, Figure 2**). AUCs were ≥0.85 at year 2, ≥0.79 at year 5, and ≥0.77 at year 6 (**Table 2, Figures S7–S8**). Unlike the results for KF, adding metabolites to benchmark models led to statistically significant AUC improvements even for Bench++ and at later prediction times, although the absolute improvements were smaller than for KF (**Table S14**). The greatest improvement was seen when plasma and urine metabolites (including unnamed) were added to the Bench+ model, yielding ΔAUCs of 0.034 (95% CI 0.014–0.057) at year 2,

0.029 (95% CI 0.017–0.040) at year 5, and 0.027 (95% CI 0.016–0.038) at year 6. Reductions in prediction error (Brier score) were most pronounced at year 6, with the largest improvement achieved by adding named plasma and urine metabolites.

## Discussion

This exploratory study demonstrates that both single metabolites and multiple-metabolite models are capable of predicting adverse kidney-related outcomes in individuals with established CKD. While we hypothesized that combining metabolites into multi-metabolite models would enhance predictive performance, the observed improvements, though statistically significant in some settings, did not reach a magnitude likely to be considered clinically meaningful.

To derive the multi-metabolite models predicting KF and CKE across various settings, we employed a component-wise boosting approach (CoxBoost) specifically designed for feature selection in high-dimensional, time-to-event data^30^. This method is well-suited for situations with many candidate predictors and can accommodate competing risk structures.

Another key advantage of CoxBoost is its ability to mandatorily include known prognostic variables, such as the KFRE variables, into the model. At each step, a single parameter is updated, using the penalized partial likelihood estimation from the previous step as offset. To prevent overfitting, the number of boosting steps (hyper-parameter) was determined in advance through 10-fold cross-validation (**Item S7**).

Across various settings, several metabolites were selected in at least two-thirds of the applicable models. Since the development of prognostic models is aimed solely at optimizing outcome prediction, the selection of metabolites does not consider their causal relationship with the outcome or any potential confounders^38^. Still, the selected metabolites may reflect information on biological meaning.

For example, plasma pseudouridine alone achieved an AUC of 0.80 (95% CI 0.77- 0.83) in predicting KF (**Table S4**). Pseudouridine has been previously linked to kidney function and CKD by several research groups (for details see Table S7 of Steinbrenner *et al.*^17^). It is a post-translationally modified nucleoside found in RNA, continuously produced and freely filtered by the kidneys. However, its net reabsorption limits its use as filtration marker^39,40^. Its connection to kidney function likely explains its selection in multi-metabolite models predicting KF only in the absence of eGFR (**Table S9**). A similar situation is observed with plasma creatinine.

Plasma N2,N5-diacetylornithine, one of the two named metabolites selected most frequently (**Figure 1**), was not identified as associated with KF or CKE in our previous project^17^. However, other studies have reported associations with eGFR, proteinuria and KF^41–43^. In contrast, the second frequently selected metabolite, urine 1-palmitoyl-2-oleoyl-GPC (16:0/18:1), was previously linked to both KF and CKE in our earlier work^17^. While serum levels of this metabolite have been associated with eGFR and proteinuria^41,43^, no associations for its urine counterpart had been described before, classifying it as a novel finding in our previous project (see Table 2 of Steinbrenner *et al*.^17^). In both instances, our observations on their prognostic value add to prior findings of their association with CKD and thus support

In a previous study, Kobayashi *et al*. had also successfully used plasma metabolites to predict inverse eGFR^44^. Of the nine metabolites selected for their model, seven were also measured in our study and three (N-acetylarginine, kynurenine and N2,N2- dimethylguanosine) were selected into some of our derived multi-metabolite models.

Overall, the derived multi-metabolite models performed well in discriminating between individuals with and without kidney disease progression and in minimizing prediction error. The performance was highest at year 2 and declined over time, a pattern also observed in the benchmark models (**Figure 2**). Prognostic models for KF consistently outperformed those for CKE, likely because KF is a more clearly defined and severe endpoint, whereas CKE includes estimated times for additional event types.

As with other proposed prognostic models, our findings reiterate that the prediction of adverse kidney-related outcomes can be improved in principle. However, the added predictive value beyond established tools like the KFRE remains too small to be clinically meaningful^9,10,36,44,45^. Even in cases where the proposed markers relevantly improve prediction, clinical implementation would depend on the ease and cost-efficiency of obtaining them. In this context, the Metabolon platform with its semi-quantitative measurements and partially unnamed metabolites is currently not suitable for routine clinical use.

One of the strengths of this study lies in the use of a well-characterized cohort with moderately decreased kidney function at baseline, comprehensive phenotyping, and long-term follow-up. Internal validation was performed whenever feasible, using resampling techniques to assess the robustness of the findings. The CoxBoost method, chosen for developing the multi-metabolite models, is particularly well suited for high-dimensional time-to-event data, such as ours. Nonetheless, we cannot rule out the possibility that alternative modeling strategies might have yielded prognostic models with better performance. Moreover, the external validation of derived models is still an open topic.

Other limitations of this project include the reliance on measurements from samples collected at a single time point (baseline). The observed decline in prognostic value over time may stem from several factors. One explanation is that metabolite levels can vary over time due to biological variability, aging, or changes in lifestyle, thereby limiting the long-term predictive utility of baseline measurements alone^46^. Supporting this, Lacruz *et al*. showed that intra-individual changes in metabolite levels over time were themselves linked to all-cause mortality^47^, suggesting the potential value of longitudinal measurements. Another important consideration is the quality of the biological samples: the samples used for metabolite profiling were collected at the baseline visit, stored at -80°C, and only analyzed several years later. It was reported that such long-term storage, even at -80°C, can affect metabolite levels^48,49^.

In summary, while we demonstrated that metabolites have prognostic value for adverse kidney-related outcomes, their contribution to improving predictive performance appears to be of limited clinical relevance. Still, our findings on the prognostic value of metabolites may inform on metabolic pathways and processes related to the progression of CKD, thereby complementing existing knowledge. Further research is warranted to further assess the role of metabolites in CKD and its progression. Importantly, the development of a clinically applicable, improved risk equation for adverse kidney outcomes will require not only the integration of multiple suggestions on novel prognostic factor models but also the incorporation of additional data sources such as longitudinal metabolite measurements, to address the limited improvements in predictive performance observed so far.

## Acknowledgement

We thank the participants in the GCKD study. The enormous effort of the study personnel at the regional centers is highly appreciated. We thank the large number of nephrologists, who have provided routine care of participants and collaborated with the GCKD study. The GCKD Investigators are listed in **Supplementary Material 1**.

## Financial Disclosure

The **GCKD** study was/is supported by the German Federal Ministry of Education and Research (Bundesministerium für Bildung und Forschung), the KfH Foundation for Preventive Medicine. Unregistered grants to support the study were provided by corporate sponsors (listed at gckd.org). Urine metabolomics were supported by Bayer Pharma AG. Plasma metabolomics has received funding from the Innovative Medicines Initiative 2 Joint Undertaking (JU) under grant agreement No 115974. The JU receives support from the European Union’s Horizon 2020 research and innovation program and EFPIA and JDRF. Any dissemination of results reflects only the author’s view; the JU is not responsible for any use that may be made of the information it contains.

The work of **AK, HB and IS** was supported by the DFG Project-ID 499552394 - SFB 1597. The work of **AK, PS, and UTS** was supported by the DFG Project-ID 431984000 - SFB 1453. The work of **EB** and **PS** was supported by the German Research Foundation (DFG, SE 2407/3-1). The work of **IS** was supported by the German Research Foundation (DFG) Project-ID 530592017 (SCHL 2292/3–1). The work of **UTS** was supported by the German Federal Ministry of Education and Research (BMBF) within the framework of the e:Med research and funding concept (grant 01ZX1912B).

## Conflict of interest

**UTS** is working for SYNLAB MVZ Humangenetik Freiburg GmbH. **ES** reports consulting fees from AstraZeneca, is an international editor of the American Journal of Kidney Diseases and a board member of the German Society of Nephrology. **KUE** reports grant support from Travere and Evotec, consulting fees from Akebia, AstraZeneca, Boeringer Ingelheim, CLS Behring, Medice, Novartis and Novonordisk, honoraria from AstraZeneca and Novonordisk, and participated on a Data Safety Monitoring Board or Advisory Board of AstraZeneca.

All other authors have nothing to disclose.

## Data availability statement

Public posting of individual level participant data is not covered by the informed patient consent form. As stated in the patient consent form and approved by the Ethics Committees, a dataset containing pseudonyms can be obtained by collaborating scientists upon approval of a scientific project proposal by the steering committee of the GCKD study: https://www.gckd.org.

## Author contributions

Research idea and study design: AK, KUE, PS Data acquisition: CB, EB, HH, IS, PO, UTS, WG Statistical analysis: EB, HB, PS Data interpretation: EB, IS, PS, UTS Drafting of manuscript: EB, PS Critical review and final approval of manuscript: AK, CB, EB, ES, HB, HH, IS, KUE, PO, PS, UTS, WG Each author contributed important intellectual content during manuscript drafting or revision and agrees to be personally accountable for the individual’s own contributions and to ensure that questions pertaining to the accuracy or integrity of any portion of the work, even one in which the author was not directly involved, are appropriately investigated and resolved, including with documentation in the literature if appropriate.

## Supplementary Material – List of content Supplementary material

Item S1: List of GCKD participating institutions and investigators

Item S2: List and definitions of baseline variables in the GCKD study

Item S3: Urine and plasma metabolite measurements in the GCKD study

Item S4: Preprocessing of metabolite measurements for statistical analysis

Item S5: Evaluation of imputation quality

Item S6: Settings considered in the analysis to develop multi-metabolite models

Item S7: Parameter settings used for the development of multi-metabolite models using CoxBoost

## Supplementary figures

Figure S1: Workflow

Figure S2: Change in AUC values and Brier scores when a single metabolite is added to a model predicting kidney failure

Figure S3: Change in AUC values and Brier scores when a single metabolite is added to a model predicting CKE

Figure S4: Illustration of the boosting algorithm for the development of a multi-metabolite model for kidney failure utilizing named plasma metabolites

Figure S5: Overview on metabolites selected for at least 8 of 12 multi-metabolite models including unnamed metabolites

Figure S6: Inclusion frequencies of metabolites according to their selection in the original model

Figure S7: Predictive performance of multi-metabolite models and respective benchmark models – outcome: kidney failure

Figure S8: Predictive performance of multi-metabolite models and respective benchmark

## Notes

### Funding Statement

The GCKD study was/is supported by the German Federal Ministry of Education and Research (Bundesministerium fuer Bildung und Forschung), the KfH Foundation for Preventive Medicine. Unregistered grants to support the study were provided by corporate sponsors (listed at gckd.org). Urine metabolomics were supported by Bayer Pharma AG. Plasma metabolomics has received funding from the Innovative Medicines Initiative 2 Joint Undertaking (JU) under grant agreement No 115974. The JU receives support from the European Union's Horizon 2020 research and innovation program and EFPIA and JDRF. Any dissemination of results reflects only the author's view; the JU is not responsible for any use that may be made of the information it contains. The work of AK, HB and IS was supported by the DFG Project-ID 499552394 - SFB 1597. The work of AK, PS, and UTS was supported by the DFG Project-ID 431984000 - SFB 1453. The work of EB and PS was supported by the German Research Foundation (DFG, SE 2407/3-1). The work of IS was supported by the German Research Foundation (DFG) Project-ID 530592017 (SCHL 2292/3-1). The work of UTS was supported by the German Federal Ministry of Education and Research (BMBF) within the framework of the e:Med research and funding concept (grant 01ZX1912B).

### Author Declarations

All ethics committees of the participating centers listed below gave ethical approval for the GCKD study and this work: Universities or Medical Faculties of Aachen, Berlin, Erlangen, Freiburg, Hannover, Heidelberg, Jena, Muenchen, Wuerzburg

## References

1. Eckardt, K.-U. et al. Evolving importance of kidney disease: from subspecialty to global health burden. Lancet Lond. Engl. 382, 158–169 (2013).

2. Jha, V. et al. Chronic kidney disease: global dimension and perspectives. Lancet Lond. Engl. 382, 260–272 (2013).

3. Pena, M. J. et al. Strategies to improve monitoring disease progression, assessing cardiovascular risk, and defining prognostic biomarkers in chronic kidney disease. Kidney Int. Suppl. 7, 107–113 (2017).

4. Kalantari, S. & Nafar, M. An update of urine and blood metabolomics in chronic kidney disease. Biomark. Med. 13, 577–597 (2019).

5. Luo, S. & Grams, M. E. Epidemiology research to foster improvement in chronic kidney disease care. Kidney Int. 97, 477–486 (2020).

6. Ramspek, C. L. et al. Kidney Failure Prediction Models: A Comprehensive External Validation Study in Patients with Advanced CKD. J. Am. Soc. Nephrol. JASN 32, 1174– 1186 (2021).

7. Tangri, N. et al. A predictive model for progression of chronic kidney disease to kidney failure. JAMA 305, 1553–1559 (2011).

8. Tangri, N. et al. Multinational Assessment of Accuracy of Equations for Predicting Risk of Kidney Failure: A Meta-analysis. JAMA 315, 164–174 (2016).

9. Schroeder, E. B. et al. Predicting 5-Year Risk of RRT in Stage 3 or 4 CKD: Development and External Validation. Clin. J. Am. Soc. Nephrol. CJASN 12, 87–94 (2017).

10. Grams, M. E. et al. Predicting timing of clinical outcomes in patients with chronic kidney disease and severely decreased glomerular filtration rate. Kidney Int. 93, 1442–1451 (2018).

11. Major, R. W. et al. The Kidney Failure Risk Equation for prediction of end stage renal disease in UK primary care: An external validation and clinical impact projection cohort study. PLoS Med. 16, e1002955 (2019).

12. Iatridi, F. et al. KDIGO 2024 Clinical Practice Guideline for the Evaluation and Management of Chronic Kidney Disease in Children and Adults: a commentary from the European Renal Best Practice (ERBP). Nephrol. Dial. Transplant. Off. Publ. Eur. Dial. Transpl. Assoc. - Eur. Ren. Assoc. 40, 273–282 (2025).

13. Hocher, B. & Adamski, J. Metabolomics for clinical use and research in chronic kidney disease. Nat. Rev. Nephrol. 13, 269–284 (2017).

14. Schultheiss, U. T., Kosch, R., Kotsis, F., Altenbuchinger, M. & Zacharias, H. U. Chronic Kidney Disease Cohort Studies: A Guide to Metabolome Analyses. Metabolites 11, 460 (2021).

15. Steinbrenner, I. et al. Urine Metabolite Levels, Adverse Kidney Outcomes, and Mortality in CKD Patients: A Metabolome-wide Association Study. Am. J. Kidney Dis. Off. J. Natl. Kidney Found. 78, 669–677.e1 (2021).

16. Steinbrenner, I. et al. Association of osteopontin with kidney function and kidney failure in chronic kidney disease patients: the GCKD study. Nephrol. Dial. Transplant. Off. Publ. Eur. Dial. Transpl. Assoc. - Eur. Ren. Assoc. 38, 1430–1438 (2023).

17. Steinbrenner, I. et al. Associations of Urine and Plasma Metabolites With Kidney Failure and Death in a Chronic Kidney Disease Cohort. Am. J. Kidney Dis. Off. J. Natl. Kidney Found. 84, 469–481 (2024).

18. Eckardt, K.-U. et al. The German Chronic Kidney Disease (GCKD) study: design and methods. Nephrol. Dial. Transplant. 27, 1454–1460 (2012).

19. Gerds, T. A., Cai, T. & Schumacher, M. The Performance of Risk Prediction Models. Biom. J. 50, 457–479 (2008).

20. Prokosch, H.-U. et al. Designing and implementing a biobanking IT framework for multiple research scenarios. Stud. Health Technol. Inform. 180, 559–563 (2012).

21. Titze, S. et al. Disease burden and risk profile in referred patients with moderate chronic kidney disease: composition of the German Chronic Kidney Disease (GCKD) cohort. Nephrol. Dial. Transplant. 30, 441–451 (2015).

22. Levey, A. S. et al. A New Equation to Estimate Glomerular Filtration Rate. Ann. Intern. Med. 150, 604 (2009).

23. Sekula, P. et al. Urine 6-Bromotryptophan: Associations with Genetic Variants and Incident End-Stage Kidney Disease. Sci. Rep. 10, 10018 (2020).

24. Sumner, L. W. et al. Proposed minimum reporting standards for chemical analysis: Chemical Analysis Working Group (CAWG) Metabolomics Standards Initiative (MSI). Metabolomics 3, 211–221 (2007).

25. Schrimpe-Rutledge, A. C., Codreanu, S. G., Sherrod, S. D. & McLean, J. A. Untargeted Metabolomics Strategies—Challenges and Emerging Directions. J. Am. Soc. Mass Spectrom. 27, 1897–1905 (2016).

26. Do, K. T. et al. Characterization of missing values in untargeted MS-based metabolomics data and evaluation of missing data handling strategies. Metabolomics 14, 128 (2018).

27. Steinbrenner, I. et al. Interactive exploration of adverse events and multimorbidity in CKD. Nephrol. Dial. Transplant. Off. Publ. Eur. Dial. Transpl. Assoc. - Eur. Ren. Assoc. 39, 2016–2024 (2024).

28. Hsu, J. Y. et al. Statistical Methods for Cohort Studies of CKD: Survival Analysis in the Setting of Competing Risks. Clin. J. Am. Soc. Nephrol. 12, 1181–1189 (2017).

29. Fine, J. P. & Gray, R. J. A Proportional Hazards Model for the Subdistribution of a Competing Risk. J. Am. Stat. Assoc. 94, 496–509 (1999).

30. Binder, H., Allignol, A., Schumacher, M. & Beyersmann, J. Boosting for high-dimensional time-to-event data with competing risks. Bioinformatics 25, 890–896 (2009).

31. Binder, H. & Schumacher, M. Adapting prediction error estimates for biased complexity selection in high-dimensional bootstrap samples. Stat. Appl. Genet. Mol. Biol. 7, Article12 (2008).

32. Gerds, T. A. & Kattan, M. W. Medical Risk Prediction: With Ties to Machine Learning. (Chapman and Hall/CRC, 2021). doi:10.1201/9781138384484.

33. Gerds, T. A., Ohlendorff, J. S. & Ozenne, B. riskRegression: Risk Regression Models and Prediction Scores for Survival Analysis with Competing Risks. 2023.12.21 10.32614/CRAN.package.riskRegression (2011).

34. Blanche, P., Dartigues, J. & Jacqmin-Gadda, H. Estimating and comparing time-dependent areas under receiver operating characteristic curves for censored event times with competing risks. Stat. Med. 32, 5381–5397 (2013).

35. Gerds, T. A. & Schumacher, M. Efron-Type Measures of Prediction Error for Survival Analysis. Biometrics 63, 1283–1287 (2007).

36. Zacharias, H. U. et al. A Predictive Model for Progression of CKD to Kidney Failure Based on Routine Laboratory Tests. Am. J. Kidney Dis. Off. J. Natl. Kidney Found. 79, 217–230.e1 (2022).

37. McShane, L. M. et al. REporting recommendations for tumour MARKer prognostic studies (REMARK). Br. J. Cancer 93, 387–391 (2005).

38. Tripepi, G., Jager, K. J., Dekker, F. W. & Zoccali, C. Testing for causality and prognosis: etiological and prognostic models. Kidney Int. 74, 1512–1515 (2008).

39. Sekula, P. et al. From Discovery to Translation: Characterization of C- Mannosyltryptophan and Pseudouridine as Markers of Kidney Function. Sci. Rep. 7, 17400 (2017).

40. Mathew, A. V. et al. Tubular dysfunction impairs renal excretion of pseudouridine in diabetic kidney disease. Am. J. Physiol. Renal Physiol. 326, F30–F38 (2024).

41. Luo, S. et al. Serum Metabolomic Alterations Associated with Proteinuria in CKD. Clin. J. Am. Soc. Nephrol. CJASN 14, 342–353 (2019).

42. Luo, S. et al. NAT8 Variants, N-Acetylated Amino Acids, and Progression of CKD. Clin. J. Am. Soc. Nephrol. CJASN 16, 37–47 (2020).

43. Lin, B. M. et al. Metabolome-wide association study of estimated glomerular filtration rates in Hispanics. Kidney Int. 101, 144–151 (2022).

44. Kobayashi, T. et al. A metabolomics-based approach for predicting stages of chronic kidney disease. Biochem. Biophys. Res. Commun. 445, 412–416 (2014).

45. Nelson, R. G. et al. Development of Risk Prediction Equations for Incident Chronic Kidney Disease. JAMA 322, 2104–2114 (2019).

46. Zeleznik, O. A. et al. Intrapersonal Stability of Plasma Metabolomic Profiles over 10 Years among Women. Metabolites 12, 372 (2022).

47. Lacruz, M. E. et al. Instability of personal human metabotype is linked to all-cause mortality. Sci. Rep. 8, 9810 (2018).

48. Haid, M. et al. Long-Term Stability of Human Plasma Metabolites during Storage at - 80 °C. J. Proteome Res. 17, 203–211 (2018).

49. Wagner-Golbs, A. et al. Effects of Long-Term Storage at -80 °C on the Human Plasma Metabolome. Metabolites 9, 99 (2019).

