## Supplementary material for "Systematic evaluation of plasma and urine metabolites to predict adverse kidney-related outcomes in chronic kidney disease: The GCKD study"

### Item S1: List of GCKD participating institutions and investigators

The nine study centers of the German Chronic Kidney Disease (GCKD) study in Germany are: RWTH Aachen University, Aachen; Charité – University-Medicine, Berlin; Friedrich-Alexander University, Erlangen; Albert-Ludwigs-University, Freiburg; Friedrich-Schiller University, Jena; Hannover Medical School, Hannover; Medical Faculty, Ruprecht-Karls University, Heidelberg; Medical Faculty, Ludwig-Maximilians-University, Munich; and Julius-Maximilians-University, Würzburg.

A list of nephrologists currently collaborating with the GCKD study is available at http://www.gckd.org.

- **University of Erlangen-Nürnberg**
  Kai-Uwe Eckardt, Heike Meiselbach, Markus P. Schneider, Mario Schiffer, Hans-Ulrich Prokosch, Barbara Bärthlein, Andreas Beck, André Reis, Arif B. Ekici, Susanne Becker, Ulrike Alberth-Schmidt, Anke Weigel, Sabine Marschall, Eugenia Schefler
- **University of Freiburg**
  Gerd Walz, Anna Köttgen, Ulla T. Schultheiß, Fruzsina Kotsis, Simone Meder, Erna Mitsch, Ursula Reinhard
- **RWTH Aachen University**
  Jürgen Floege, Turgay Saritas, Alice Groß
- **Charité, University Medicine Berlin**
  Elke Schaeffner, Seema Baid-Agrawal, Kerstin Theisen
- **Hannover Medical School**
  Hermann Haller
- **University of Heidelberg**
  Martin Zeier, Claudia Sommerer, Mehtap Aykac
- **University of Jena**
  Gunter Wolf, Martin Busch, Rainer Paul
- **Ludwig-Maximilians University of München**
  Thomas Sitter
- **University of Würzburg**
  Christoph Wanner, Vera Krane, Antje Börner-Klein, Britta Bauer
- **Medical University of Innsbruck, Division of Genetic Epidemiology**
  Florian Kronenberg, Julia Raschenberger, Barbara Kollerits, Lukas Forer, Sebastian Schönherr, Hansi Weissensteiner
- **University of Regensburg, Institute of Functional Genomics**
  Peter J. Oefner, Wolfram Gronwald
- **Institute of Medical Biometry, Informatics and Epidemiology, Medical Faculty, University of Bonn**

Matthias Schmid, Jennifer Nadal

### Item S2: List and definitions of baseline variables in the GCKD study

| **Variable** | **Comment** | **Missing values**  **(N=5,217)** |
| --- | --- | --- |
| **Clinical information used in both prediction models MET+ and MET++ (KFRE factors)** | | |
| Age | in years | 0 (0%) |
| Sex | self-reported; male (0), female (1) | 0 (0%) |
| eGFR | estimated glomerular filtration rate; in mL/min/1.73m²; eGFR was estimated using the Chronic Kidney Disease Epidemiology Collaboration (CKD-EPI) formula;^1^ an IDMS traceable enzymatic assay (Creatinine Plus, Roche) was used to measure serum creatinine (mg/dL). | 55 (1%) |
| UACR | urinary albumin-to-creatinine ratio; in mg/g; UACR was based on urinary creatinine (IDMS traceable enzymatic assay, Creatinine Plus, Roche; mg/dL) and urinary albumin (ALBU-XS assay (Roche/Hitachi Diagnostics GmbH, Mannheim, Germany; mg/L); for statistical analysis, UACR was ln-transformed (natural logarithm). | 90 (2%) |
| **Further clinical information used in prediction model MET++** | | |
| smoking | self-reported; non-smoker (0), ex-smoker (1), smoker (2); individuals reporting current daily or occasional smoking were classified as smokers. | 16 (0%) |
| BMI | body mass index; in kg/m²; BMI was calculated from individual’s body weight and height, and corrected for any amputations. | 54 (1%) |
| BP,  systolic | blood pressure, systolic; in mmHg; systolic BP was calculated as the mean out of three measurements spaced one minute apart after five minutes resting using a standardized device (OMRON M5 Professional, Mannheim, Germany). | 34 (1%) |
| serum  albumin | in g/L; serum albumin was quantified using a turbidimetric method (Tina-quant, Roche, Germany). | 57 (1%) |
| CRP | high-sensitivity C-reactive protein; in mg/L; CRP was measured using an immunoturbidimetric test (CRPHS, Roche, Germany) on a Roche/Hitachi MODULAR (P); for statistical analysis, CRP was ln-transformed (natural logarithm). | 56 (1%) |
| total  cholesterol | in mg/dL; total cholesterol was measured using an enzymatic colorimetric method (CHOD-PAP, Roche, Germany) on a Roche/Hitachi MODULAR (P). | 62 (1%) |
| diabetes | presence of diabetes at baseline; no (0) or yes (1); diabetes was defined as either a baseline serum hemoglobin A1c (HbA1c) of ≥6.5% or documented use of diabetes medication (Anatomical Therapeutic Chemical [ATC] code beginning with ‘A10’;^2^). | 0 (0%) |
| CHD | prevalent coronary heart disease; no (0) or yes (1); CHD was defined as self-reported history of myocardial infarction, bypass operation, or percutaneous coronary intervention.^3^ | 2 (0%) |
| stroke | self-reported history of stroke; no (0) or yes (1) | 2 (0%) |
| loop  diuretics | self-reported intake of blood pressure-lowering medications: loop diuretics; no (0) or yes (1); group of loop diuretics was defined based on ATC codes (ATC ‘C03C’). | 0 (0%) |

Abbreviations: **KFRE:** kidney failure risk equation; **MET+**: metabolite model including major prognostic variables for kidney failure; **MET++**: metabolite model including major prognostic variables for kidney failure and further reported prognostic variables of adverse kidney-related outcomes

### Item S3: Urine and plasma metabolite measurements in the GCKD study

Spot urine and plasma samples collected at baseline from participants of the German Chronic Kidney Disease (GCKD) study were used to determine metabolite concentrations using non-targeted liquid chromatography-mass spectrometry (LC-MS) analysis provided by Metabolon, Inc. (Durham, NC, USA)^4,5^. While plasma samples were measured in one batch (2020), urine samples had been analysed in three batches (2016-2017).

Sample preparation, LC-MS, and metabolite identification were described previously^6,7^. Briefly, four different UPLC-MS/MS methods were used for maximum coverage of hydrophilic and hydrophobic molecules by positive and negative ion mode electrospray ionization (range: 70-1000 *m/z* [mass-to-charge ratio])^8^. To identify metabolites, ion features in the GCKD study samples were compared to a reference library using an in-house software^9,10^. The Metabolon’s reference library contains information on molecular mass, retention time, preferred adducts, in-source fragments and associated MS spectra of various chemical standards and is continuously curated. Levels of the metabolites were measured as area under the integrals of respective peaks^10^. Raw area counts were normalized to account for inter-day instrument variation by the median value for each run-day and represent semi-quantitative (unit-less) measurements. To align metabolite identification between assessed urine and plasma samples from the GCKD study, metabolite identification in processed urine samples was repeated at the same time plasma sample results were processed.

While some identified metabolites belong to different super-pathways (e.g., amino acids, xenobiotics), a large proportion of metabolites is still only partially characterized or unnamed (i.e., of unknown structural identity). Unless a named metabolite is marked by an asterisk, named metabolites conform to highest confidence level (level 1) of the Metabolomics Standards Initiative^11,12^.

### Item S4: Preprocessing of metabolite measurements for statistical analysis

After transfer of measurements to the Institute of Genetic Epidemiology (Medical Center – University of Freiburg, Germany), an in-house pipeline was used for data quality control and preparation of data for statistical analysis.

Details on quality control checks can be found elsewhere^6,7^. The quality-controlled datasets contain semi-quantitative measurements of 1,513 urine metabolites in 5,088 of 5,217 GCKD study participants (97.5%) and 1,416 plasma metabolites in 5,144 of 5,217 GCKD study participants (98.6%). Semi-quantitative levels show different degrees of completeness (range: 0-99.9%).

In preparation of the statistical analysis, urine measurements were normalized to harmonize differences in dilution using the probabilistic quotient method^13^. As in previous studies^6,14,15^, all metabolite measurements were log_2_-transformed to reduce skewness in data.

Furthermore, non-xenobiotics with <50% missing levels and xenobiotics with <5% missing levels were imputed per matrix. Using *k*-nearest neighbor (*knn*) algorithm with k=10, the levels of 1,096 plasma metabolites and 1,129 urine metabolites were imputed^16^. Annotation of imputed plasma and urine metabolites are listed in **Supplementary Tables 1** and **2**.

### Item S5: Evaluation of imputation quality

To assess whether the imputation of missing measurements affects analyses, two sensitivity analyses were conducted:

1. **Question:** does the imputation affect the correlation structure of metabolite pairs within the same matrix?

For this purpose, Spearman correlation coefficients of metabolite pairs per matrix were calculated from unimputed and imputed data. Respective coefficients for a given pair are plotted against each other in the figure:

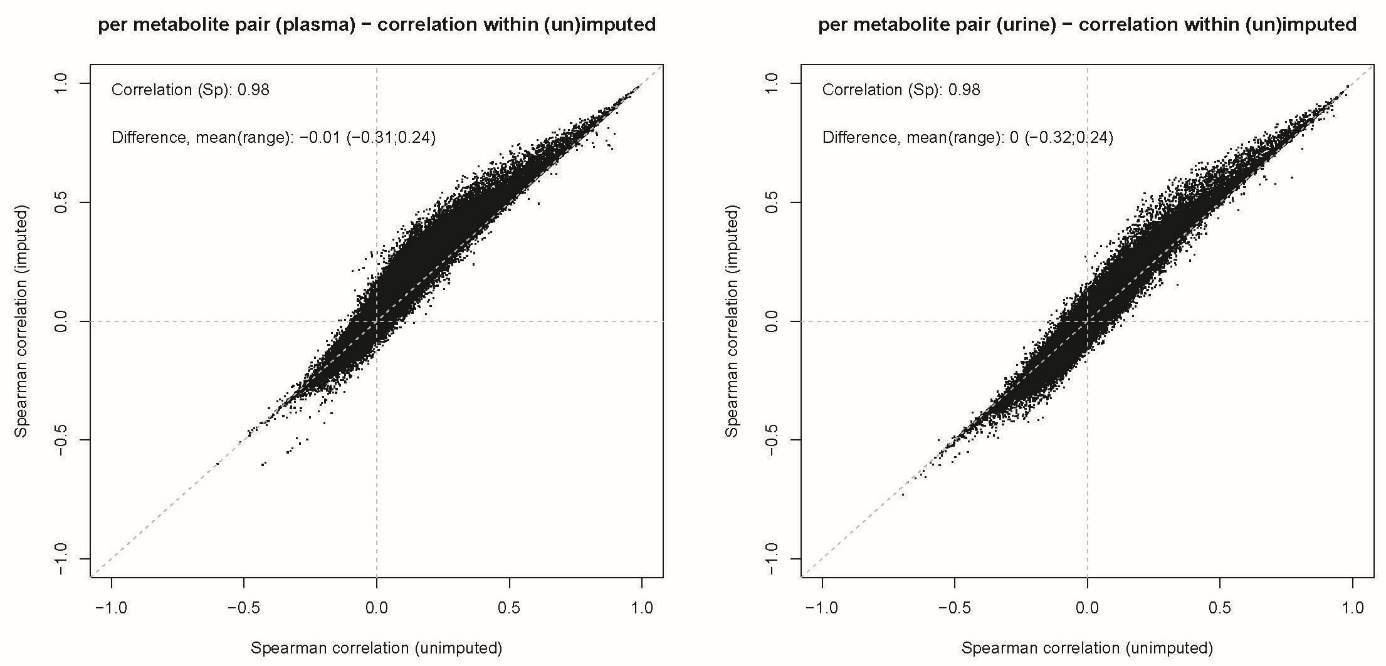

For each pair of metabolites within each matrix, the Spearman correlation coefficient was calculated with unimputed (x-axis) and imputed (y-axis) data and displayed against each other. Overall, the imputation retained general correlation structure that was seen before imputation. left: for plasma metabolites, right: for urine metabolites.

**Conclusion:** Imputation of missing values essentially retains correlation structure.

2. **Question:** does the imputation affect the direction and significance of associations?

For this purpose, the association analysis as reported in Steinbrenner et al. was repeated using the imputed data^17^. The table shows Spearman correlation coefficients for effect estimates (p-values) based on the results from each stage and setting:

| **Matrix** | | **Plasma** | | **Urine** | |
| --- | --- | --- | --- | --- | --- |
| **Outcome** | | **KF** | **CKE** | **KF** | **CKE** |
| **Analysis stage** | **Discovery** | 0.98 (0.95) | 0.99 (0.96) | 0.97 (0.92) | 0.98 (0.94) |
|  | **Replication** | 0.98 (0.95) | 0.98 (0.95) | 0.96 (0.91) | 0.98 (0.94) |
|  | **Meta-analysis** | 0.99 (0.95) | 0.99 (0.96) | 0.97 (0.93) | 0.98 (0.94) |

For illustration, the scatter plots display obtained effect estimates (beta) and -log10 p-values obtained from the meta-analyses:

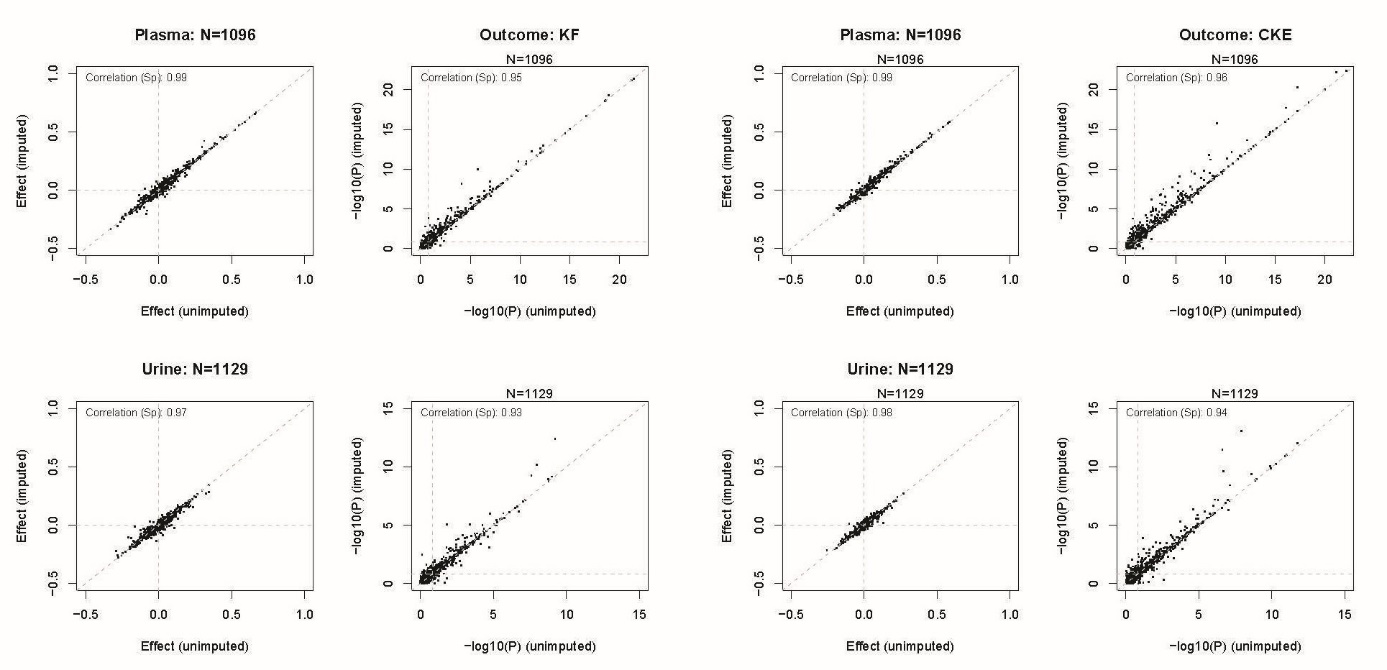

Effect estimates (adjusted for age, sex, eGFR, ln-transformed UACR) and –log10 p-values from the analysis of kidney failure (KF) are presented on the left side and from analysis of composite kidney endpoint (CKD) on the right side. The upper row refers to the analysis of plasma metabolites and lower row to the analysis of urinary metabolites. In addition, the Spearman correlation coefficient is displayed. Overall, association results align very well.

Conclusion: Overall, results obtained from the analysis of imputed data show good agreement with those of unimputed data.

### Item S6: Settings considered in the analysis to develop multi-metabolite models

To systematically assess the potential of plasma and urinary metabolites, several settings for the development of multi-metabolite prediction models were considered:

| **Setting No** | **Metabolites** | **Outcome** | **Matrix** | **Model** |
| --- | --- | --- | --- | --- |
| 1 | Only named metabolites | Kidney failure  (KF) | Plasma | MET |
| 2 |  |  |  | MET+ |
| 3 |  |  |  | MET++ |
| 4 |  |  | Urine | MET |
| 5 |  |  |  | MET+ |
| 6 |  |  |  | MET++ |
| 7 |  |  | Plasma + Urine | MET |
| 8 |  |  |  | MET+ |
| 9 |  |  |  | MET++ |
| 10 |  | Composite  kidney endpoint  (CKE) | Plasma | MET |
| 11 |  |  |  | MET+ |
| 12 |  |  |  | MET++ |
| 13 |  |  | Urine | MET |
| 14 |  |  |  | MET+ |
| 15 |  |  |  | MET++ |
| 16 |  |  | Plasma + Urine | MET |
| 17 |  |  |  | MET+ |
| 18 |  |  |  | MET++ |
| 19 | All metabolites | Kidney failure  (KF) | Plasma | MET |
| 20 |  |  |  | MET+ |
| 21 |  |  |  | MET++ |
| 22 |  |  | Urine | MET |
| 23 |  |  |  | MET+ |
| 24 |  |  |  | MET++ |
| 25 |  |  | Plasma + Urine | MET |
| 26 |  |  |  | MET+ |
| 27 |  |  |  | MET++ |
| 28 |  | Composite  kidney endpoint  (CKE) | Plasma | MET |
| 29 |  |  |  | MET+ |
| 30 |  |  |  | MET++ |
| 31 |  |  | Urine | MET |
| 32 |  |  |  | MET+ |
| 33 |  |  |  | MET++ |
| 34 |  |  | Plasma + Urine | MET |
| 35 |  |  |  | MET+ |
| 36 |  |  |  | MET++ |

**MET:** model including only metabolite information; **MET+:** model including metabolite information and the four variables from the Kidney Failure Risk Equation (age, sex, eGFR, and ln-transformed UACR); **MET++:** model including metabolite information and an extended set of prognostic factors (see **Item S2** for details)

### Item S7: Parameter settings used for the development of multi-metabolite models using CoxBoost

For the development of multi-metabolite models, penalized regression was utilized using the component-wise boosting algorithm *CoxBoost* (v1.5, <https://github.com/binderh/CoxBoost>)^18^. A 2-step approach was used (i) to determine the number of necessary boosting steps for a given dataset and (ii) to conduct feature selection with the determined number of steps.

**List of chosen parameters:**

1. Determination of number of necessary boosting steps using the function *cv.CoxBoost*

   input:
   - vector with time to event/censoring information
   - vector with event indicator
   - matrix with exposure information

parameters:
- maxstepno = 500 (in stability assessment: 750)
- K = 10
- type = ”verweij”
- penalty = 9*number of events of interest
- *for models with clinical information*: unpen.index = list of numbers indicating
 columns in exposure matrix containing clinical information

2. Selection of metabolites using the function *CoxBoost*

input: *as above*

parameters:
- stepno = number of necessary boosting steps determined in step 1
- penalty = 9*number of events of interest (default)

- criterion = "score"

- cmprsk = "sh"

- standardize = FALSE

- *for models with clinical information*: unpen.index = list of numbers indicating
 columns in exposure matrix containing clinical information

#### SUPPLEMENTARY FIGURES

### Figure S1: Workflow

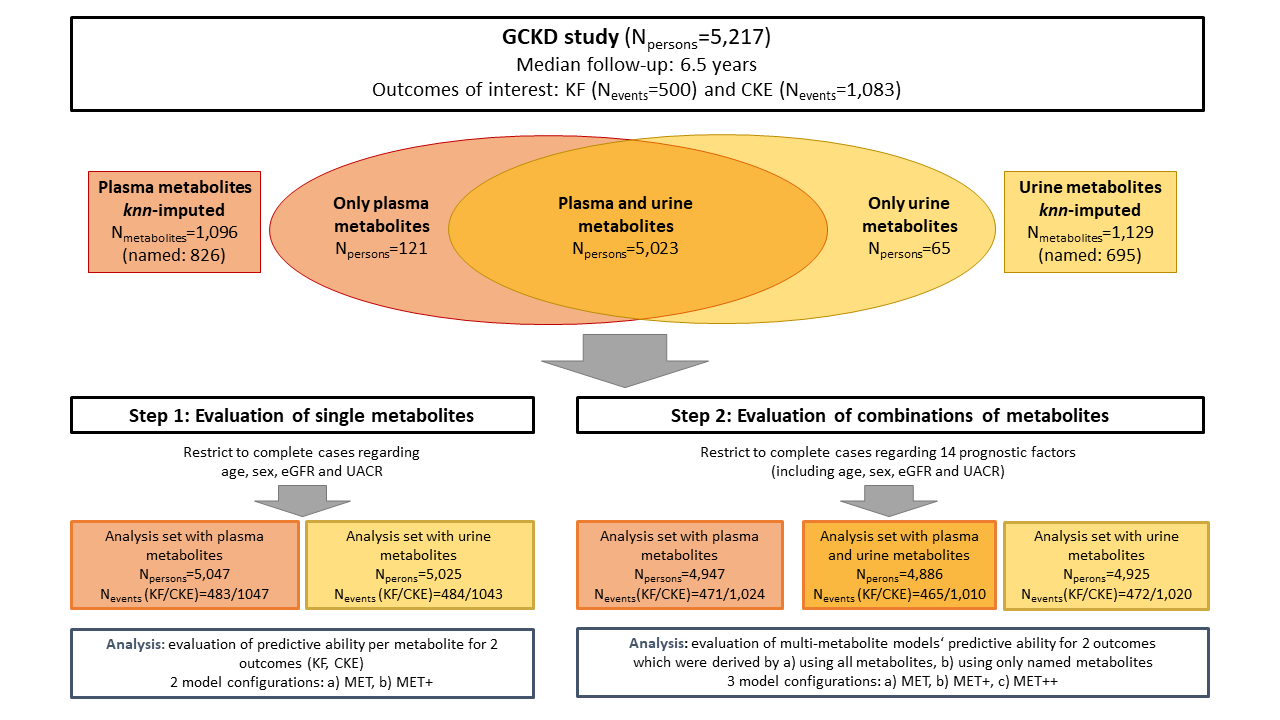

See **Item S3** and **Item S4** for details on measure-ments of metabolites and their preparations for the analyses; See **Item S2** for details on 14 prognostic factors considered in the analyses.

Abbreviations:
**KF:** kidney failure
**CKE:** composite kidney endpoint.

In total, plasma measurements available for 5,144 persons and urine for 5,088 (overlap N=5,023). For a total of 8 persons no measurements were available at all.

### Figure S2: Change in AUC values and Brier scores when a single metabolite is added to a model predicting kidney failure

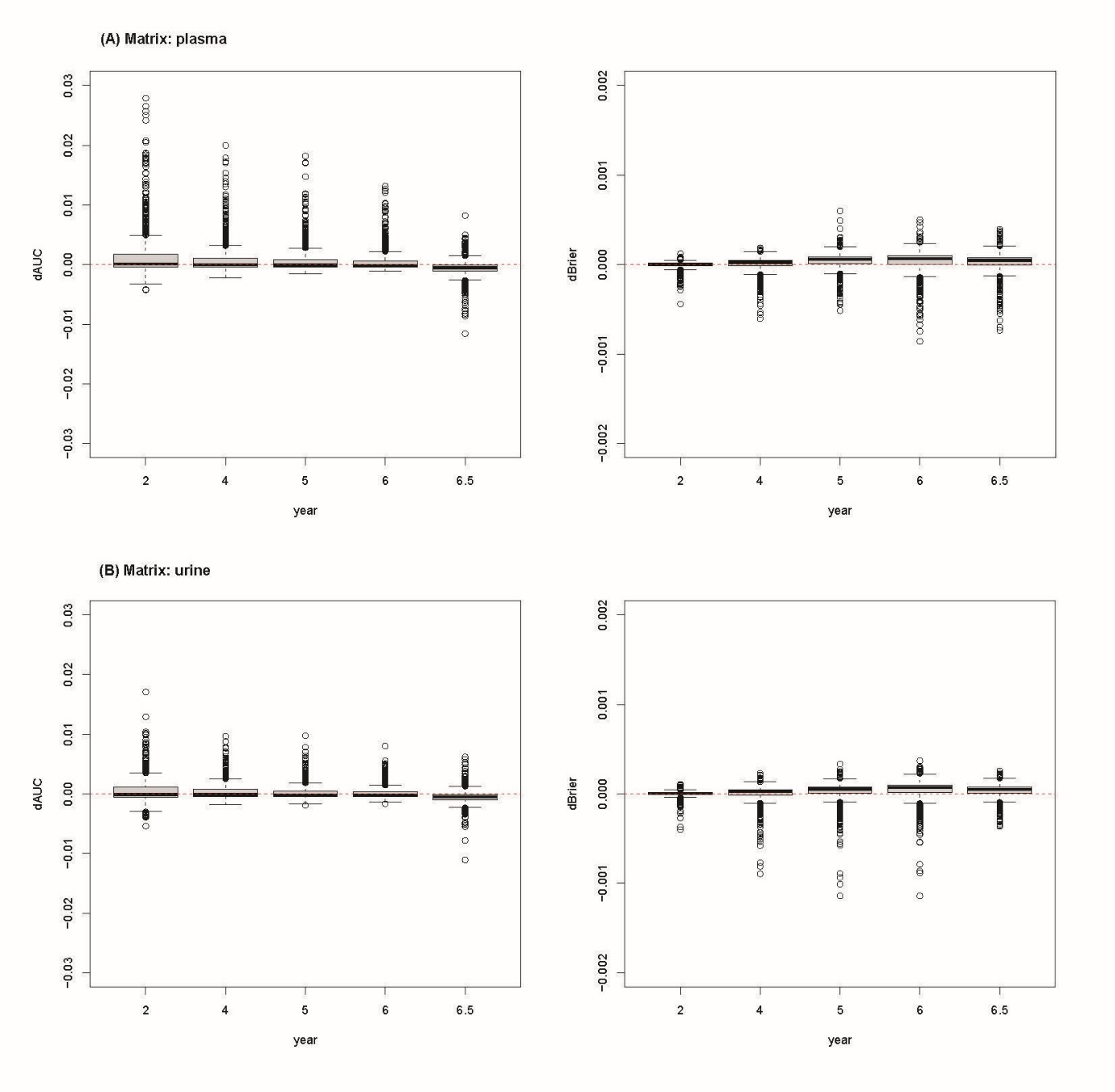

Change in AUC (dAUC) and Brier score (dBrier) is measured as the difference between the AUC or Brier score from the model with four KFRE variables (age, sex, eGFR, and ln-transformed UACR) and the model that additionally includes a single metabolite. Positive values of dAUC reflect improvement when adding the metabolite. For Brier score, negative values reflect improvement. The plot shows the box plots for dAUC (left) and dBrier (right) at specific times per matrix.

**Conclusion:** While metabolites do not improve the prediction of the respective outcome on average, there are several instances where metabolites can improve the prediction (dAUC >0 or dBrier <0).

Abbreviations: **AUC:** area under the receiver operating characteristic curve; **KFRE:** kidney failure risk equation; **eGFR:** estimated glomerular filtration rate; **UACR:** urinary albumin-to-creatinine ratio.

### Figure S3: Change in AUC values and Brier scores when a single metabolite is added to a model predicting CKE

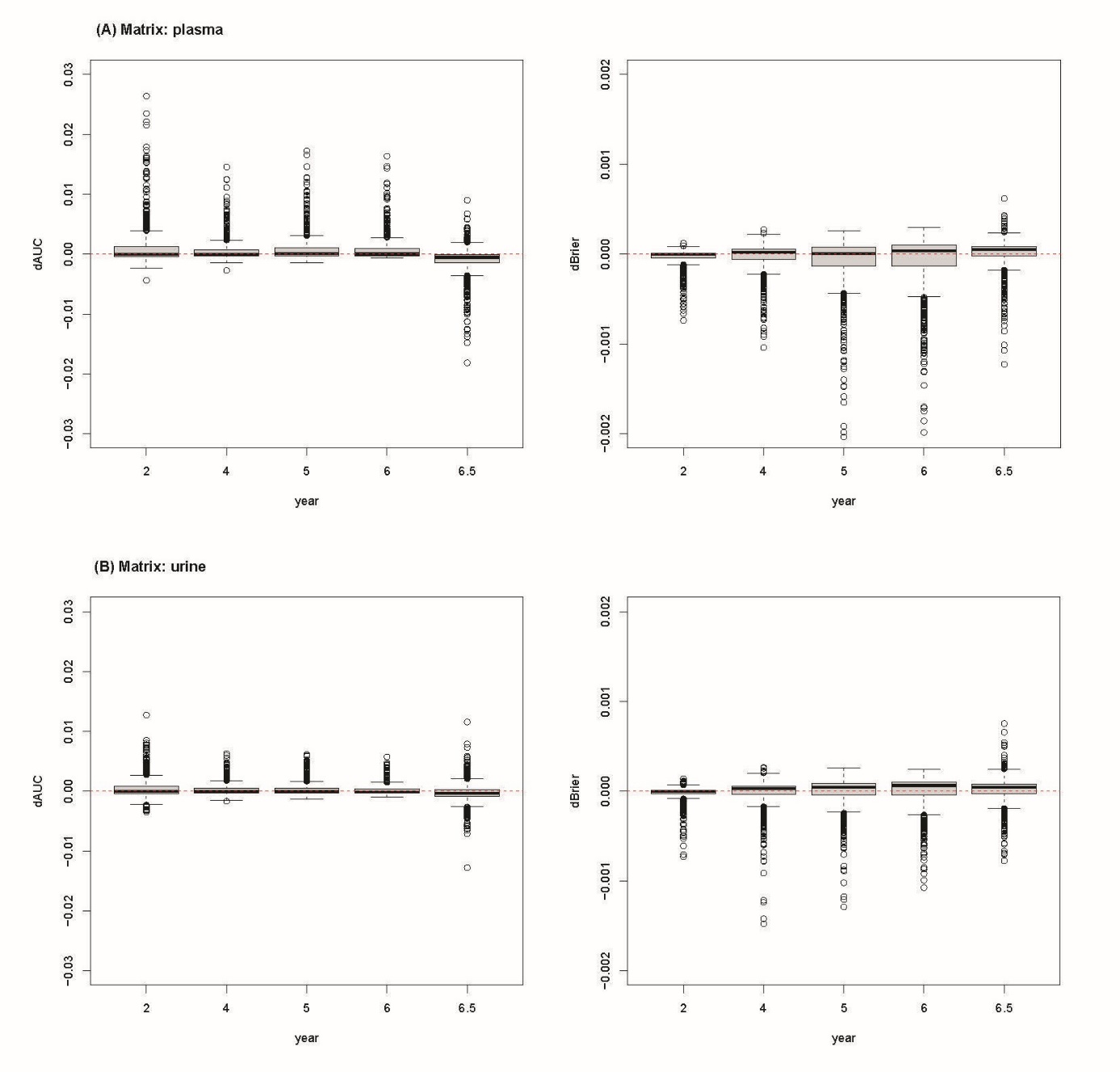

Change in AUC (dAUC) and Brier score (dBrier) is measured as the difference between the AUC or Brier score from the model with four KFRE variables (age, sex, eGFR, and ln-transformed UACR) and the model that additionally includes a single metabolite. Positive values of dAUC reflect improvement when adding the metabolite. For Brier score, negative values reflect improvement. The plot shows the box plots for dAUC (left) and dBrier (right) at specific times per matrix.

**Conclusion:** While metabolites do not improve the prediction of the respective outcome on average, there are several instances where metabolites can improve the prediction (dAUC >0 or dBrier <0).

Abbreviations: **AUC:** area under the receiver operating characteristic curve; **KFRE:** kidney failure risk equation; **eGFR:** estimated glomerular filtration rate; **UACR:** urinary albumin-to-creatinine ratio.

### Figure S4: Illustration of the boosting algorithm for the development of a multi-metabolite model for kidney failure utilizing named plasma metabolites

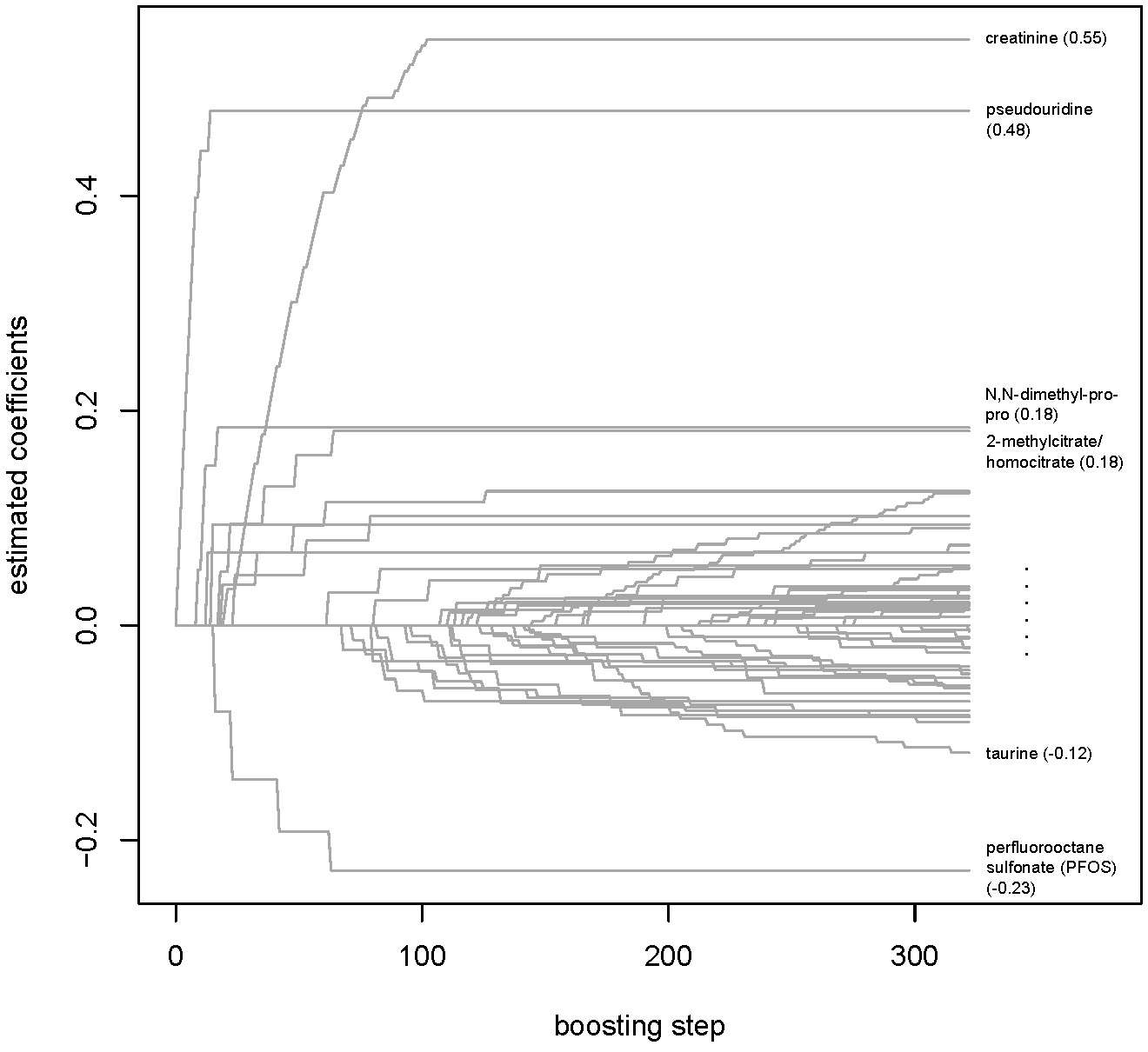

For the development of multi-metabolite models, penalized regression was utilized using the component-wise boosting algorithm CoxBoost (v1.5, <https://github.com/binderh/CoxBoost>)^18^. In a first step, the optimal number of boosting step is determined using 10-fold cross-validation. In the here presented setting (outcome: kidney failure, model: metabolites only [MET], matrix: named plasma metabolites), the determined number of boosting step was 322. In a second step, the selection of metabolites was done using the predefined number of steps. At each step, the coefficient of one variable is updated and any metabolite with a non-zero coefficient is selected into the model.

This Figure shows the coefficient paths for this specific setting. Selected paths are annotated with the names of the respective metabolite and its final coefficient in brackets. In the end, 50 metabolites had been seleted.

See Item S7 for details on parameters used in the call of CoxBoost and Table S9 for selected metabolites.

### Figure S5: Overview on metabolites selected for at least 8 of 12 multi-metabolite models including unnamed metabolites

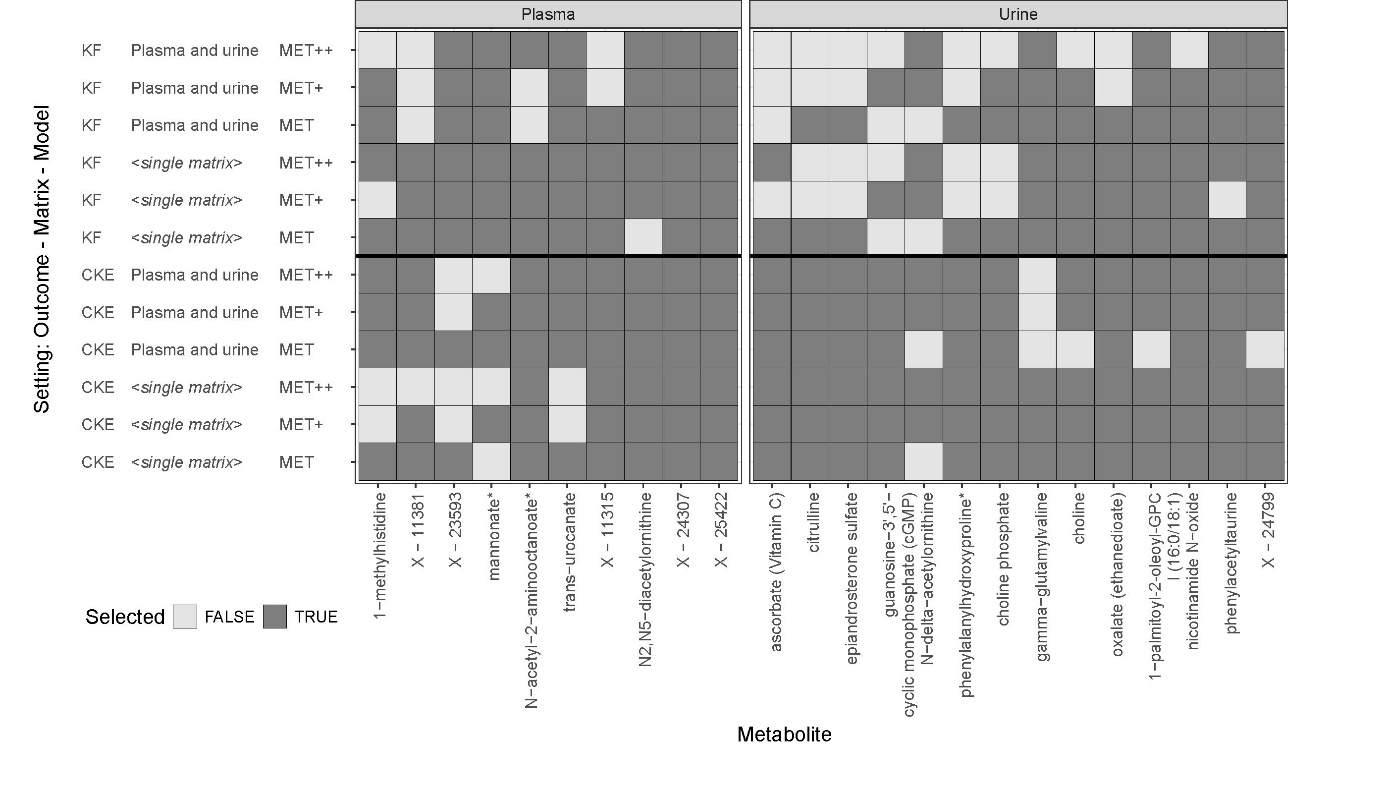

Multi-metabolite models including unnamed metabolites were derived under 18 different settings. Each metabolite measured in plasma or in urine, can be selected in maximum 12 of the 18 settings. This heatmap highlights the metabolites chosen in at least 2/3 of the settings (N=8).

Abbreviations:
**KF:** kidney failure
**CKE:** composite kidney endpoint
**MET:** model incorporating only metabolite information
**MET+:** metabolite model including metabolite information and the four variables from the Kidney Failure Risk Equation (age, sex, eGFR, and ln-transformed UACR)
**MET++:** metabolite model including metabolite information and an extended set of prognostic factors (see **Item S2** for details)

### Figure S6: Inclusion frequencies of metabolites according to their selection in the original model

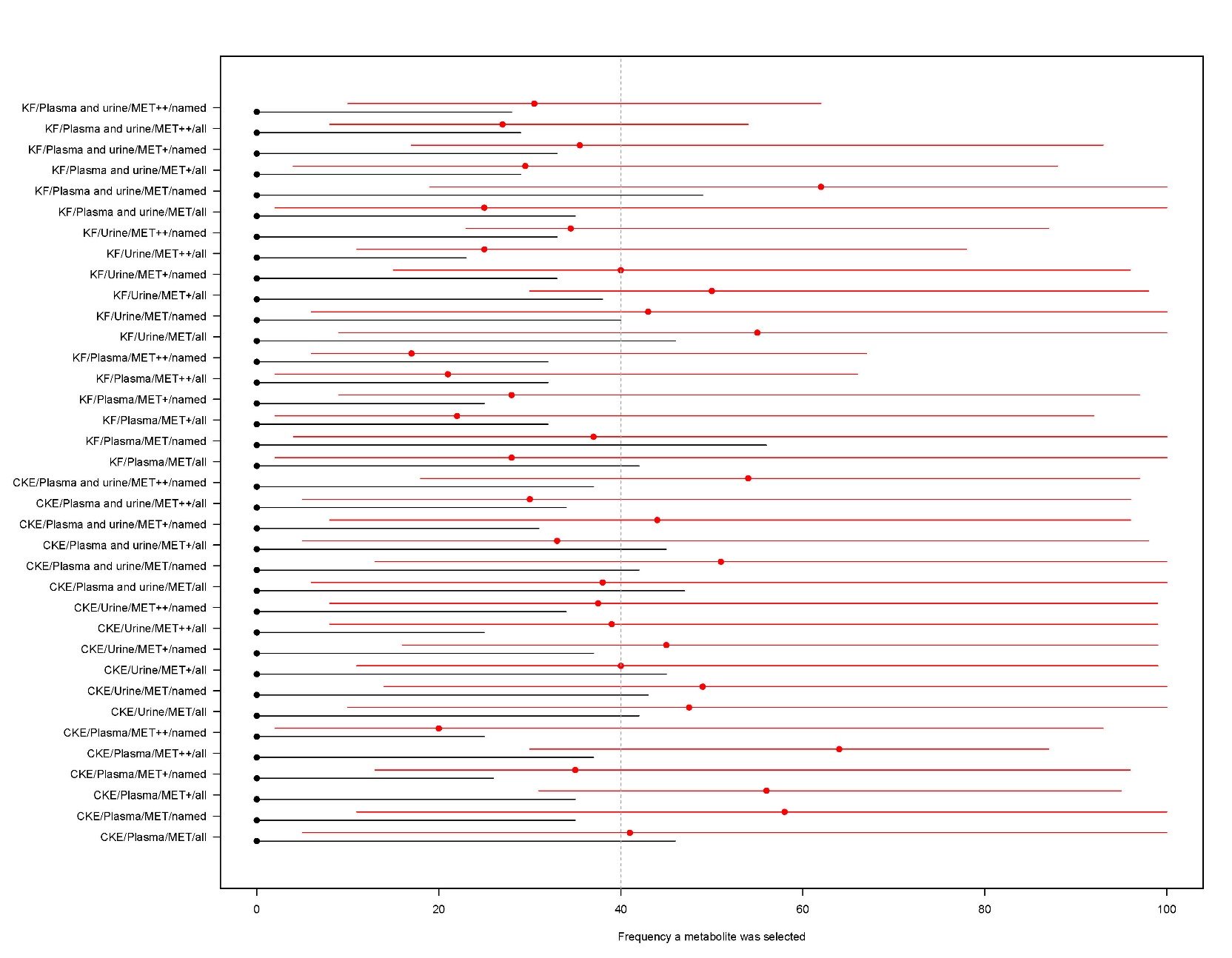

For each of 36 settings, the distribution of metabolite selection frequency across 100 bootstrap samples is shown (point: median, line: minimum and maximum). Metabolites were categorized into a group of metabolites that were selected originally in the respective setting (red) and that were not selected originally (black).

The grey vertical line represents a threshold at 40%.

**Conclusion:** Metabolites that were selected into the model originally, were also selected more often in bootstrap samples.

### Figure S7: Predictive performance of multi-metabolite models and respective benchmark models – outcome: kidney failure

(A) Matrix: plasma, named	(B) Matrix: plasma, all

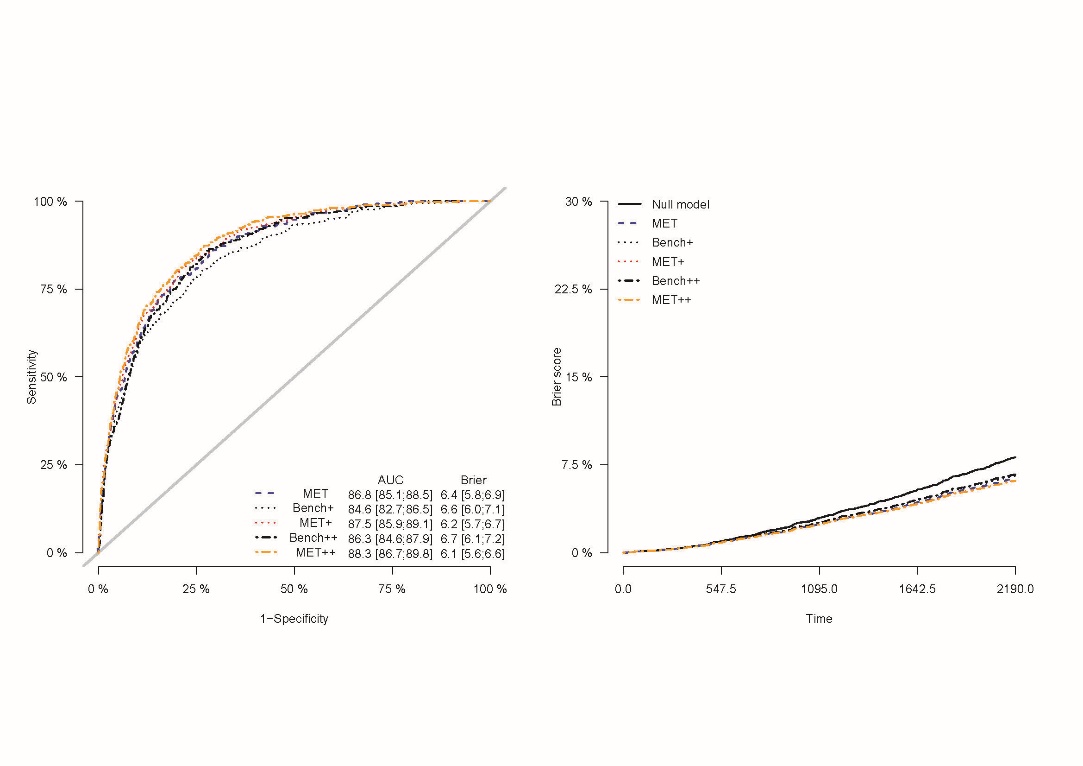

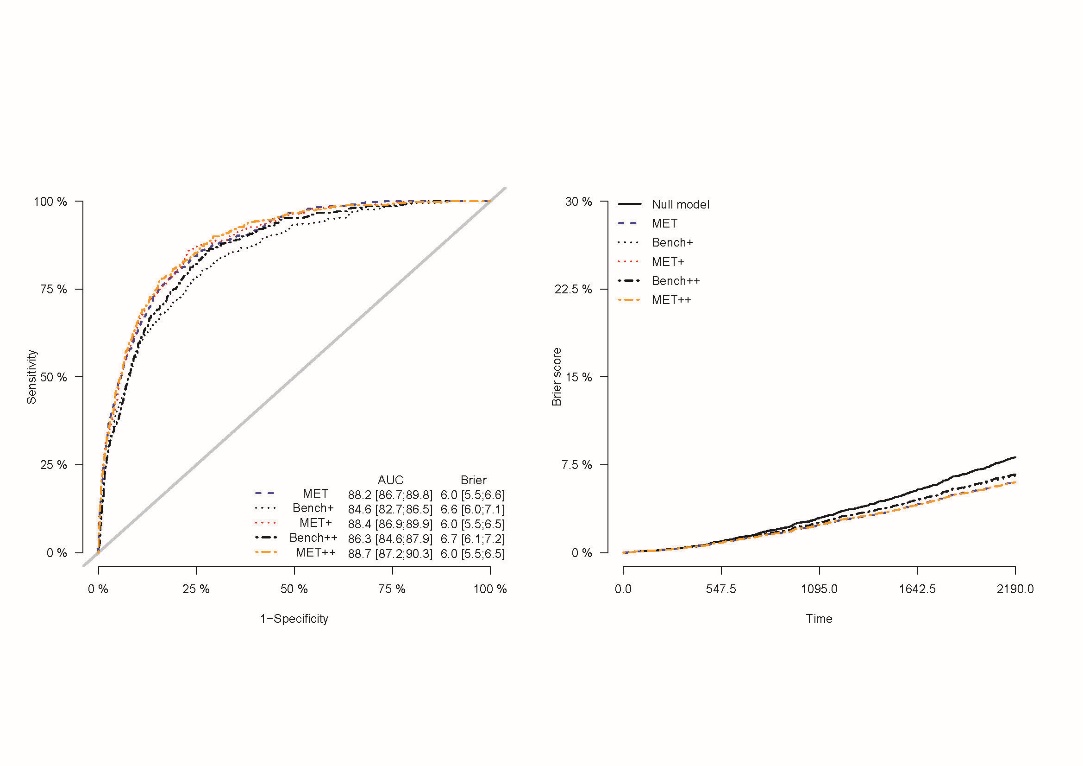

Left: receiver operating characteristics curve at time = 6 years; Right: Prediction error curve

**Null model:** model without any explanatory variable; **MET:** metabolite model**; Bench+:** model only including major prognostic variables for kidney failure; **MET+**: metabolite model including major prognostic variables for kidney failure; **Bench++:** model only including major prognostic variables for kidney failure and further reported prognostic variables of adverse kidney-related outcomes; **MET++**: metabolite model including major prognostic variables for kidney failure and further reported prognostic variables of adverse kidney-related outcomes; See **Item S2** for details on prognostic variables included.

**Figure S7** … *continue*

(C) Matrix: urine, named	(D) Matrix: urine, all

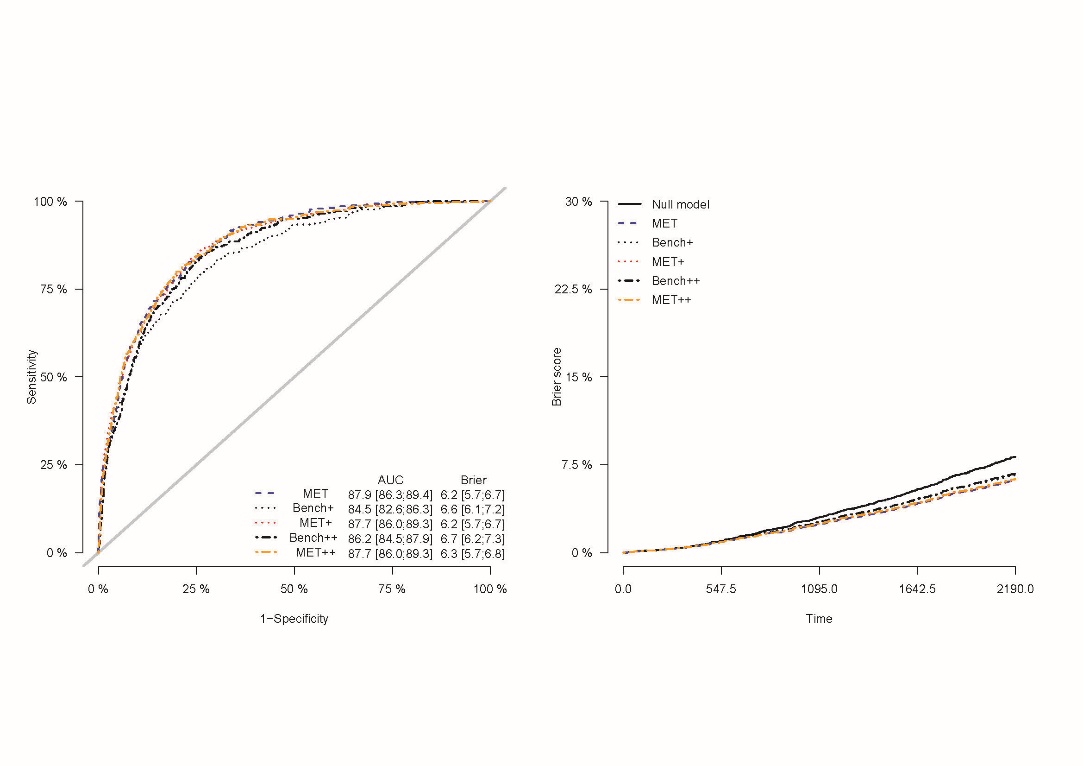

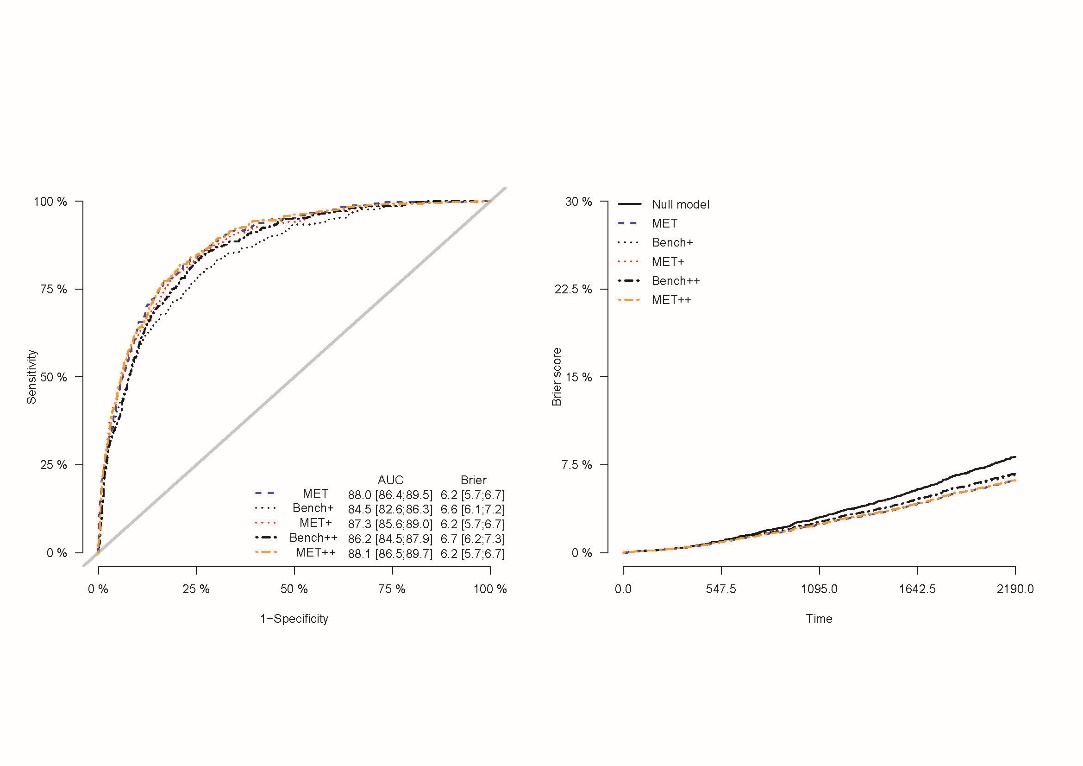

Left: receiver operating characteristics curve at time = 6 years; Right: Prediction error curve

**Null model:** model without any explanatory variable; **MET:** metabolite model**; Bench+:** model only including major prognostic variables for kidney failure; **MET+**: metabolite model including major prognostic variables for kidney failure; **Bench++:** model only including major prognostic variables for kidney failure and further reported prognostic variables of adverse kidney-related outcomes; **MET++**: metabolite model including major prognostic variables for kidney failure and further reported prognostic variables of adverse kidney-related outcomes; See **Item S2** for details on prognostic variables included.

**Figure S7** … *continue*

(E) Matrix: plasma and urine, named	(F) Matrix: plasma and urine, all

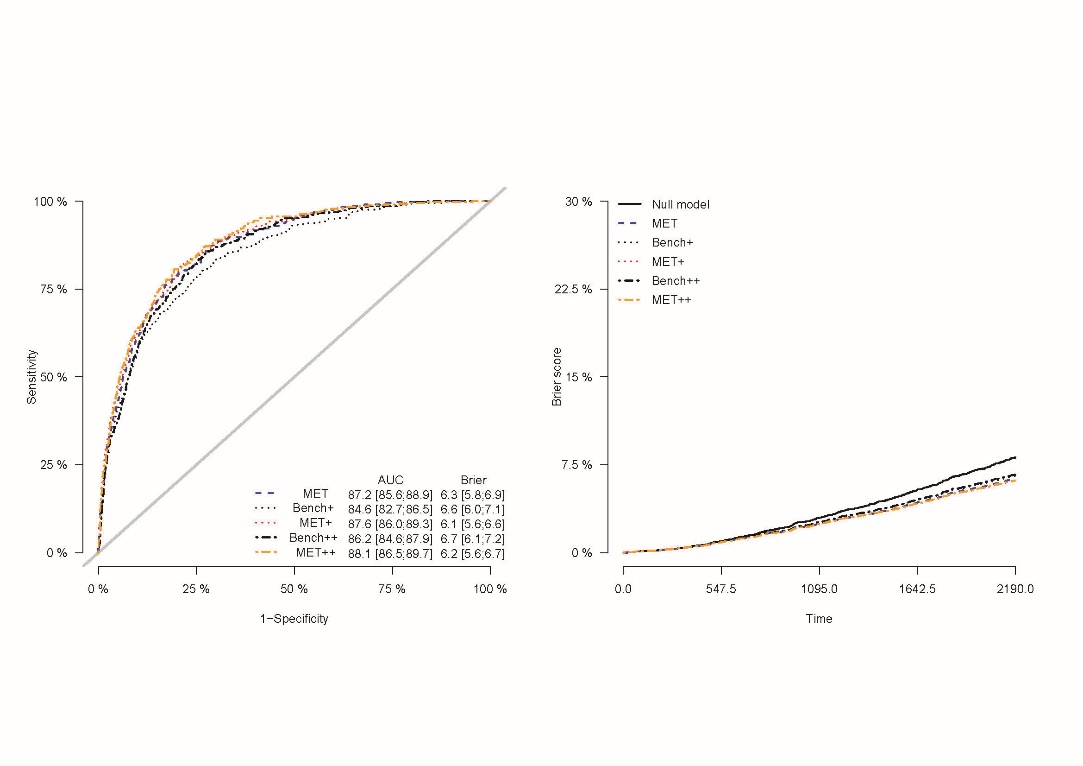

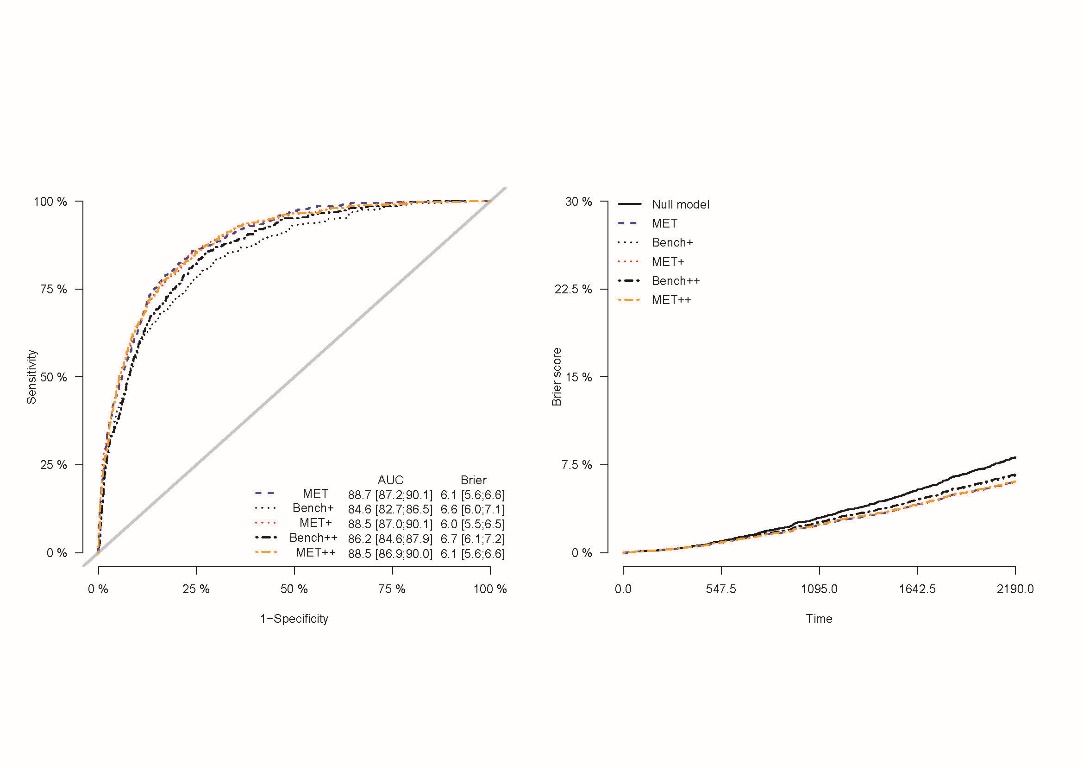

Left: receiver operating characteristics curve at time = 6 years; Right: Prediction error curve

**Null model:** model without any explanatory variable; **MET:** metabolite model**; Bench+:** model only including major prognostic variables for kidney failure; **MET+**: metabolite model including major prognostic variables for kidney failure; **Bench++:** model only including major prognostic variables for kidney failure and further reported prognostic variables of adverse kidney-related outcomes; **MET++**: metabolite model including major prognostic variables for kidney failure and further reported prognostic variables of adverse kidney-related outcomes; See **Item S2** for details on prognostic variables included.

### Figure S8: Predictive performance of multi-metabolite models and respective benchmark models – outcome: composite kidney endpoint

(A) Matrix: plasma, named	(B) Matrix: plasma, all

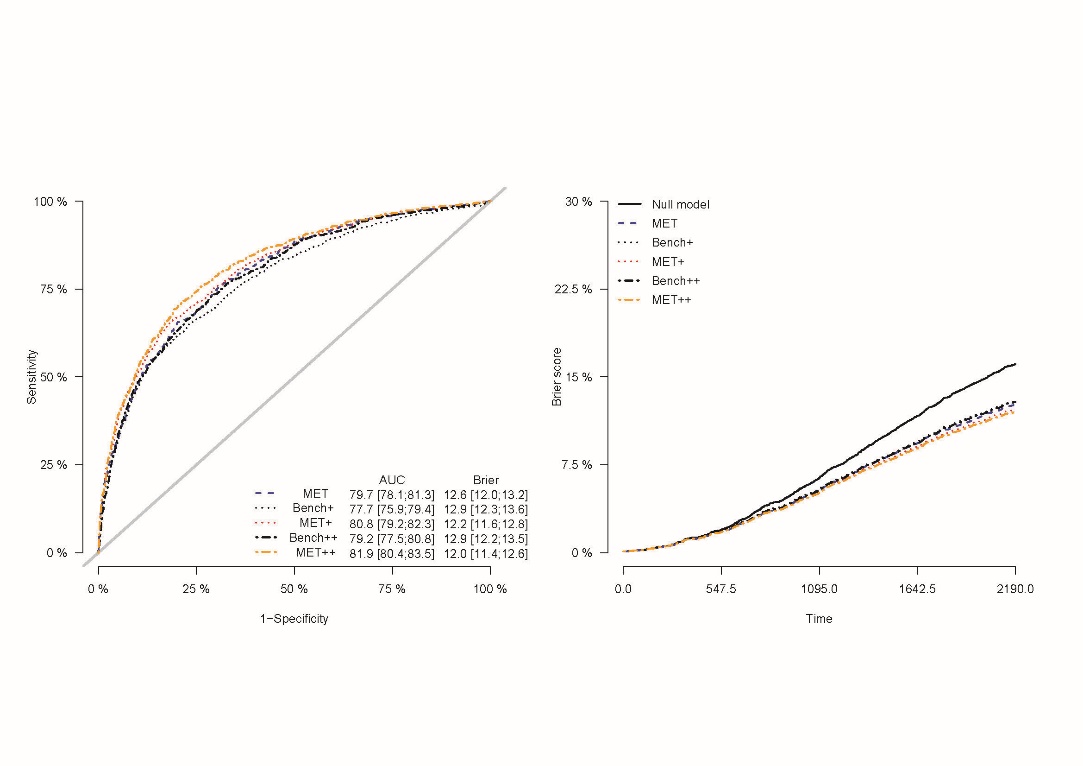

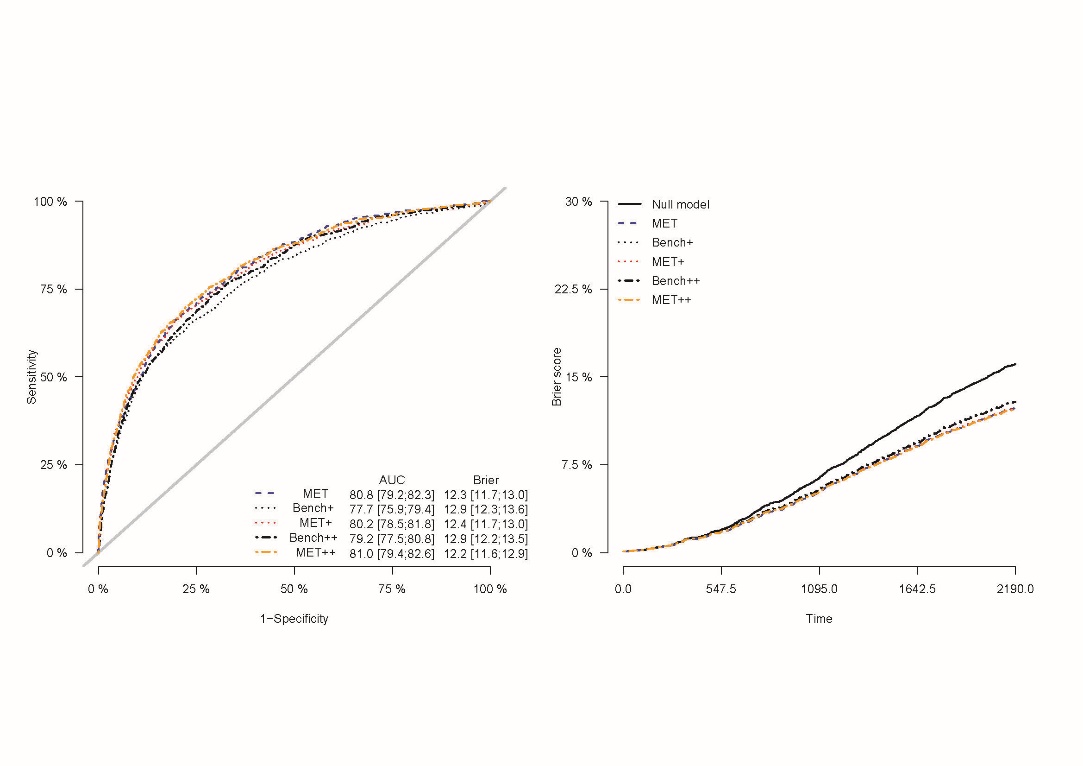

Left: receiver operating characteristics curve at time = 6 years; Right: Prediction error curve

**Null model:** model without any explanatory variable; **MET:** metabolite model**; Bench+:** model only including major prognostic variables for kidney failure; **MET+**: metabolite model including major prognostic variables for kidney failure; **Bench++:** model only including major prognostic variables for kidney failure and further reported prognostic variables of adverse kidney-related outcomes; **MET++**: metabolite model including major prognostic variables for kidney failure and further reported prognostic variables of adverse kidney-related outcomes; See **Item S2** for details on prognostic variables included.

**Figure S8** … *continue*

(C) Matrix: urine, named	(D) Matrix: urine, all

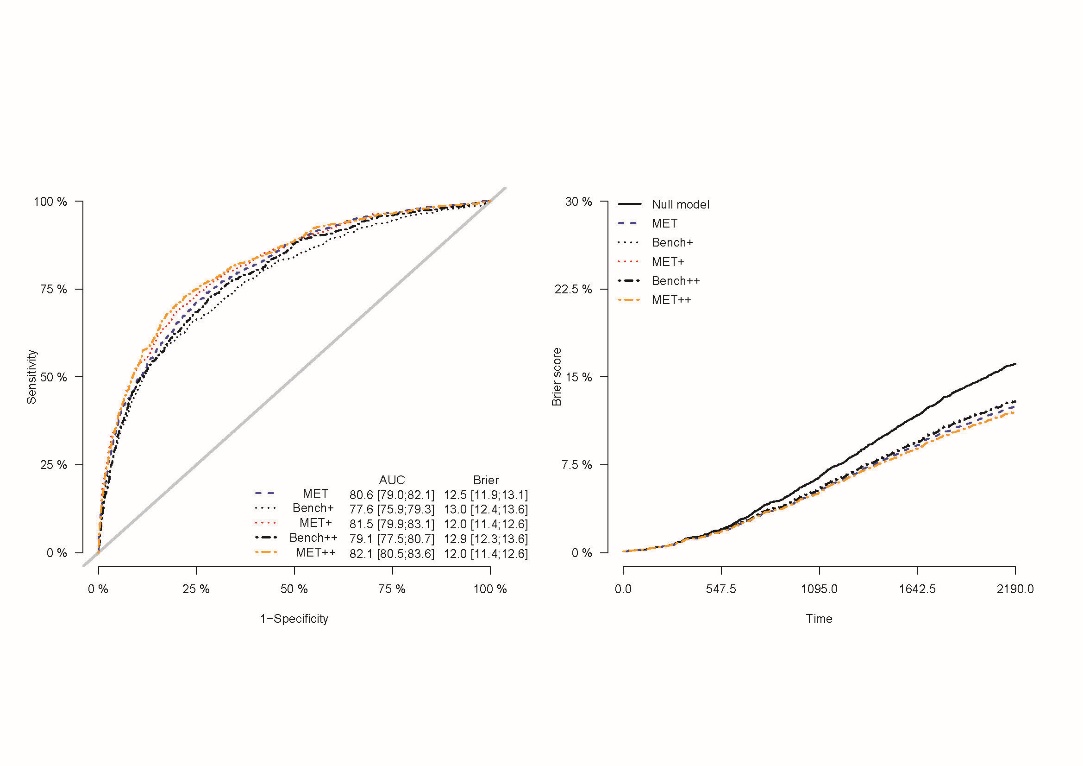

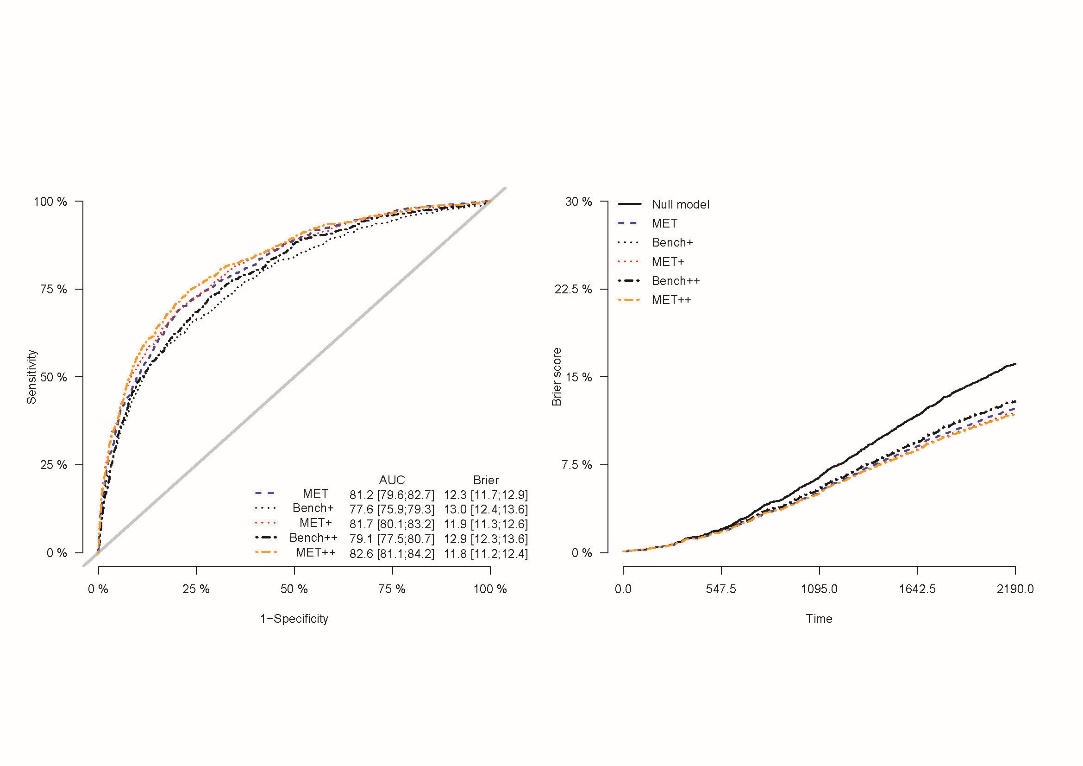

Left: receiver operating characteristics curve at time = 6 years; Right: Prediction error curve

**Null model:** model without any explanatory variable; **MET:** metabolite model**; Bench+:** model only including major prognostic variables for kidney failure; **MET+**: metabolite model including major prognostic variables for kidney failure; **Bench++:** model only including major prognostic variables for kidney failure and further reported prognostic variables of adverse kidney-related outcomes; **MET++**: metabolite model including major prognostic variables for kidney failure and further reported prognostic variables of adverse kidney-related outcomes; See **Item S2** for details on prognostic variables included.

**Figure S8** … *continue*

(E) Matrix: plasma and urine, named	(F) Matrix: plasma and urine, all

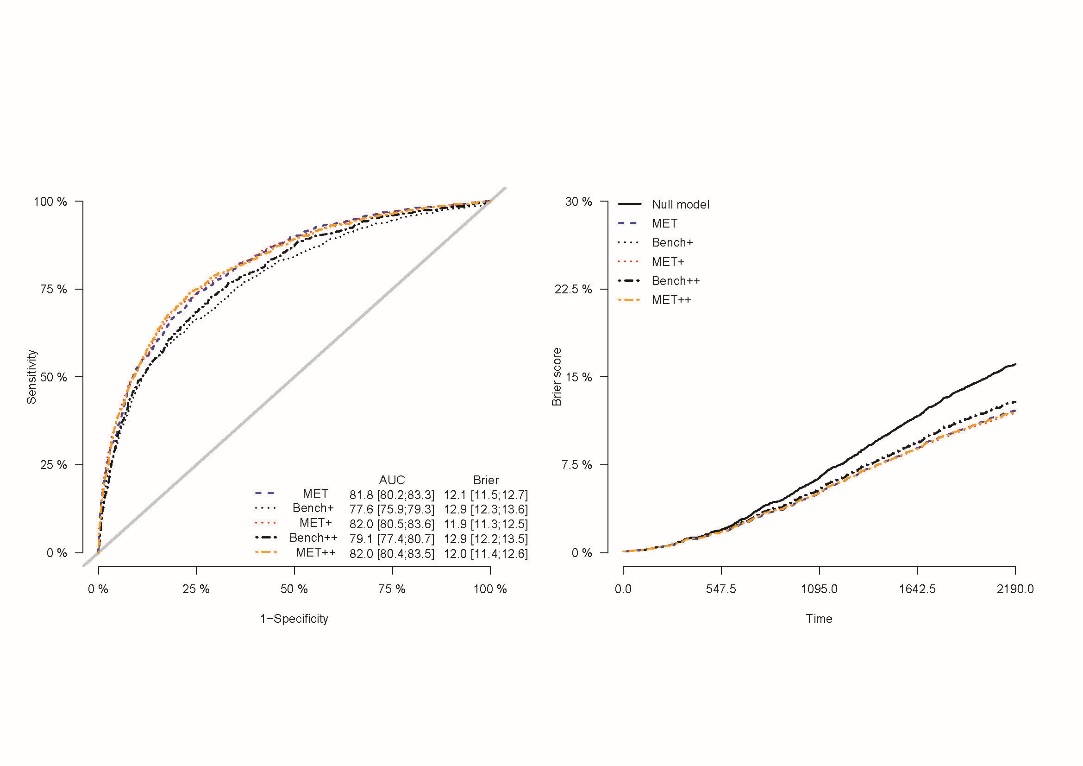

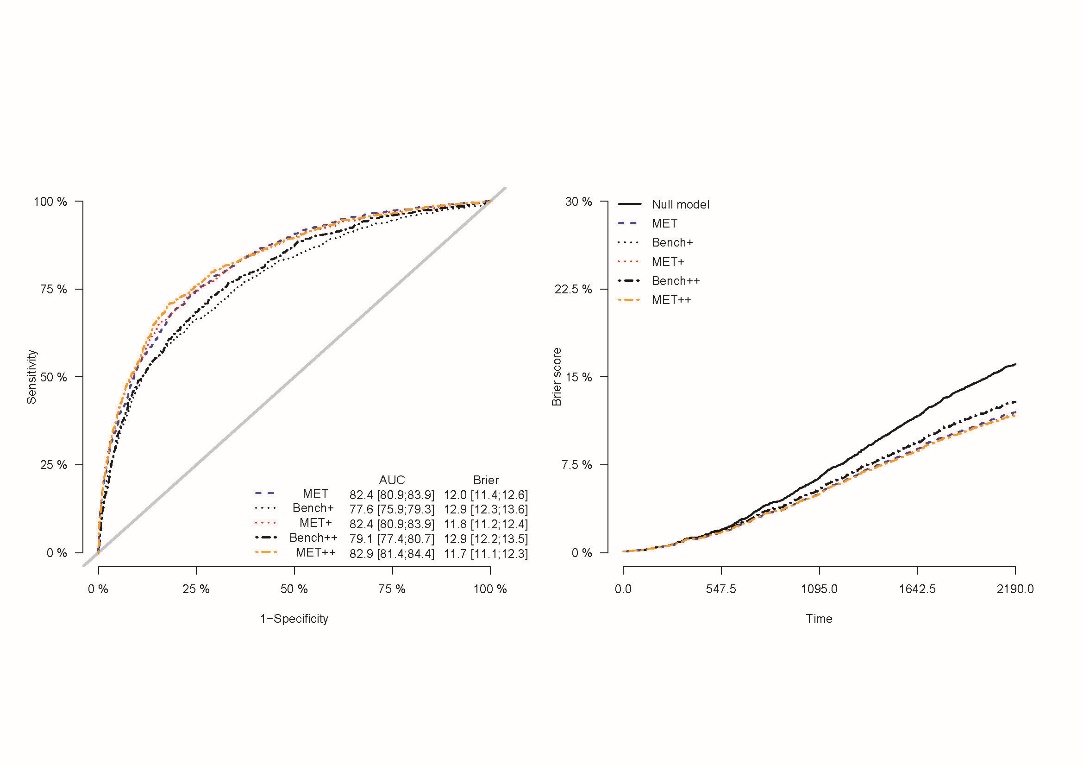

Left: receiver operating characteristics curve at time = 6 years; Right: Prediction error curve

**Null model:** model without any explanatory variable; **MET:** metabolite model**; Bench+:** model only including major prognostic variables for kidney failure; **MET+**: metabolite model including major prognostic variables for kidney failure; **Bench++:** model only including major prognostic variables for kidney failure and further reported prognostic variables of adverse kidney-related outcomes; **MET++**: metabolite model including major prognostic variables for kidney failure and further reported prognostic variables of adverse kidney-related outcomes; See **Item S2** for details on prognostic variables included.

#### SUPPLEMENTARY TABLES

*See separate excel file*

#### REFERENCES

1.	Levey AS, Stevens LA, Schmid CH, et al. A new equation to estimate glomerular filtration rate. *Ann Intern Med*. 2009;150(9):604-612. doi:10.7326/0003-4819-150-9-200905050-00006

2.	Schmidt IM, Hübner S, Nadal J, et al. Patterns of medication use and the burden of polypharmacy in patients with chronic kidney disease: the German Chronic Kidney Disease study. *Clin Kidney J*. 2019;12(5):663-672. doi:10.1093/ckj/sfz046

3.	Titze S, Schmid M, Köttgen A, et al. Disease burden and risk profile in referred patients with moderate chronic kidney disease: composition of the German Chronic Kidney Disease (GCKD) cohort. *Nephrol Dial Transplant*. 2015;30(3):441-451. doi:10.1093/ndt/gfu294

4.	Eckardt KU, Bärthlein B, Baid-Agrawal S, et al. The German Chronic Kidney Disease (GCKD) study: design and methods. *Nephrol Dial Transplant*. 2012;27(4):1454-1460. doi:10.1093/ndt/gfr456

5.	Prokosch HU, Mate S, Christoph J, et al. Designing and implementing a biobanking IT framework for multiple research scenarios. *Stud Health Technol Inform*. 2012;180:559-563.

6.	Schlosser P, Li Y, Sekula P, et al. Genetic studies of urinary metabolites illuminate mechanisms of detoxification and excretion in humans. *Nat Genet*. 2020;52(2):167-176. doi:10.1038/s41588-019-0567-8

7.	Sekula P, Tin A, Schultheiss UT, et al. Urine 6-Bromotryptophan: Associations with Genetic Variants and Incident End-Stage Kidney Disease. *Sci Rep*. 2020;10(1):10018. doi:10.1038/s41598-020-66334-w

8.	Shin SY, Fauman EB, Petersen AK, et al. An atlas of genetic influences on human blood metabolites. *Nat Genet*. 2014;46(6):543-550. doi:10.1038/ng.2982

9.	Dehaven CD, Evans AM, Dai H, Lawton KA. Organization of GC/MS and LC/MS metabolomics data into chemical libraries. *J Cheminformatics*. 2010;2(1):9. doi:10.1186/1758-2946-2-9

10.	Jump RLP, Polinkovsky A, Hurless K, et al. Metabolomics analysis identifies intestinal microbiota-derived biomarkers of colonization resistance in clindamycin-treated mice. *PloS One*. 2014;9(7):e101267. doi:10.1371/journal.pone.0101267

11.	Sumner LW, Amberg A, Barrett D, et al. Proposed minimum reporting standards for chemical analysis Chemical Analysis Working Group (CAWG) Metabolomics Standards Initiative (MSI). *Metabolomics Off J Metabolomic Soc*. 2007;3(3):211-221. doi:10.1007/s11306-007-0082-2

12.	Schrimpe-Rutledge AC, Codreanu SG, Sherrod SD, McLean JA. Untargeted Metabolomics Strategies-Challenges and Emerging Directions. *J Am Soc Mass Spectrom*. 2016;27(12):1897-1905. doi:10.1007/s13361-016-1469-y

13.	Dieterle F, Ross A, Schlotterbeck G, Senn H. Probabilistic quotient normalization as robust method to account for dilution of complex biological mixtures. Application in 1H NMR metabonomics. *Anal Chem*. 2006;78(13):4281-4290. doi:10.1021/ac051632c

14.	Steinbrenner I, Schultheiss UT, Kotsis F, et al. Urine Metabolite Levels, Adverse Kidney Outcomes, and Mortality in CKD Patients: A Metabolome-wide Association Study. *Am J Kidney Dis Off J Natl Kidney Found*. 2021;78(5):669-677.e1. doi:10.1053/j.ajkd.2021.01.018

15.	Schlosser P, Scherer N, Grundner-Culemann F, et al. Genetic studies of paired metabolomes reveal enzymatic and transport processes at the interface of plasma and urine. *Nat Genet*. 2023;55(6):995-1008. doi:10.1038/s41588-023-01409-8

16.	Do KT, Wahl S, Raffler J, et al. Characterization of missing values in untargeted MS-based metabolomics data and evaluation of missing data handling strategies. *Metabolomics Off J Metabolomic Soc*. 2018;14(10):128. doi:10.1007/s11306-018-1420-2

17.	Steinbrenner I, Schultheiss UT, Bächle H, et al. Associations of Urine and Plasma Metabolites With Kidney Failure and Death in a Chronic Kidney Disease Cohort. *Am J Kidney Dis Off J Natl Kidney Found*. 2024;84(4):469-481. doi:10.1053/j.ajkd.2024.03.028

18.	Binder H, Allignol A, Schumacher M, Beyersmann J. Boosting for high-dimensional time-to-event data with competing risks. *Bioinformatics*. 2009;25(7):890-896. doi:10.1093/bioinformatics/btp088
